# Characterising the effects of genetic liability to autoimmune conditions on pregnancy outcomes using Mendelian Randomization

**DOI:** 10.1101/2025.08.12.25333479

**Authors:** Elisabeth Aiton, Nancy S McBride, Gemma L Clayton, Ana Goncalves Soares, Tom Bond, Qian Yang, Charikleia Chatzigeorgiou, Jane West, Benjamin G Faber, Katherine Birchenall, Christy Burden, Maria C Magnus, Deborah A Lawlor, Maria Carolina Borges

**Affiliations:** Population Health Sciences, Bristol Medical School, University of Bristol, UK; MRC Integrative Epidemiology Unit at the University of Bristol, Bristol, UK; Department of Endocrine and Metabolic Diseases, Shanghai Institute of Endocrine and Metabolic Diseases, Ruijin Hospital, Shanghai Jiao Tong University School of Medicine, Shanghai, China; Shanghai National Clinical Research Center for Metabolic Diseases, Key Laboratory for Endocrine and Metabolic Diseases of the National Health Commission of the PR China, Shanghai Key Laboratory for Endocrine Tumor, Lifecycle Health Management Center, Ruijin Hospital, Shanghai Jiao Tong University School of Medicine, Shanghai, China; School of Medicine and Population Health, University of Sheffield, UK; Musculoskeletal Research Unit, University of Bristol, UK; Department of Obstetrics and Gynaecology, St Michael’s Hospital, Bristol, UK; Translational Health Sciences, Bristol Medical School, University of Bristol, Bristol, UK; School of Medicine and Public Health, Hunter Medical Research Institute, University of Newcastle, Australia; Centre for Fertility and Health, Norwegian Institute of Public Health, Oslo, Norway

**Keywords:** Autoimmune conditions, causality, genetic, Mendelian randomization, pregnancy

## Abstract

**Background:** Autoimmune conditions are common in women of reproductive age. They are associated with increased risk of adverse pregnancy outcomes; whether this is causal is unclear. Our aim was to explore the effects of autoimmune condition liability on pregnancy outcomes.

**Methods:** We conducted two-sample Mendelian randomization (MR) to estimate effects of liability to ten autoimmune conditions on nine primary and seven secondary pregnancy outcomes. We used data from the MR-PREG collaboration including up to 934,566 pregnancies. Main analyses used the inverse variance weighted method; sensitivity analyses were used to explore bias due to pleiotropic variants and fetal genetics.

**Findings:** We found evidence for 14 effects of autoimmune condition liability on primary pregnancy outcomes that were robust across sensitivity analyses. Higher liability to Hashimoto’s thyroiditis was protective against large for gestational age and increased risks of hypertensive disorders of pregnancy (HDP), preterm birth (PTB), and neonatal intensive care unit (NICU) admission. For instance, risk of PTB increased by 6% (OR=1.06 (95%CI: 1.02, 1.11)) per doubling in log odds of Hashimoto’s thyroiditis. Liability to type 1 diabetes increased risks of HDP and NICU admission, as well as gestational diabetes mellitus (GDM) and stillbirth.

Liability to rheumatoid arthritis increased risks of NICU admission, HDP, and GDM. Higher ankylosing spondylitis liability reduced risks of HDP and increased risks of low Apgar score, while systemic lupus erythematosus liability increased risks of PTB only. For multiple sclerosis, systemic sclerosis, coeliac disease, inflammatory bowel disease, and psoriasis, we did not detect any robust effects of increased liability.

**Interpretation:** We observed higher liability to Hashimoto’s thyroiditis, type 1 diabetes, and rheumatic conditions cause increased risks of adverse pregnancy outcomes, suggesting the need for enhanced antenatal monitoring of women with these conditions.

**Funding:** Wellcome Trust, UK Medical Research Council

## Introduction

Autoimmune conditions are increasing in incidence globally and are more common in women. In Europe, one in eight women is affected(^1^), and many are diagnosed before or during their reproductive years(^1^). Pregnancy itself results in marked changes to the immune system which are essential to maintain tolerance to the semi-allogeneic fetus while remaining resilient to infections.

There is a complex interplay between autoimmune conditions and pregnancy. On one hand, pregnancy can have a profound effect on the activity of autoimmune conditions. For example, patients with multiple sclerosis typically experience fewer relapses during pregnancy(^2^). On the other hand, pregnant women with autoimmune conditions are often at greater risk of developing adverse pregnancy outcomes in observational studies(^3^). Rheumatic conditions, encompassing both systemic lupus erythematosus and rheumatoid arthritis, have been associated with increased risks of stillbirth, preeclampsia, and preterm birth (PTB)(^3^). Clinical hypothyroidism, most often caused by Hashimoto’s thyroiditis, is associated with higher risks of miscarriage, hypertensive disorders of pregnancy (HDP), and PTB(^3,4^). Type 1 diabetes associates with a substantially increased risk of stillbirth and PTB(^3^), and inflammatory bowel disease is also associated with PTB(^3^) and admission to neonatal intensive care unit (NICU)(^5^).

Even in systematic reviews with meta-analyses of observational studies, the associations between autoimmune conditions and pregnancy outcomes are often underpowered for rare outcomes such as stillbirth, low Apgar score and admission to NICU(^3^). These observational associations may not be causal and could be explained by residual confounding due to inadequate adjustment for maternal characteristics, such as socioeconomic position, high body mass index and comorbid autoimmune conditions. Research in this area is limited, and little evidence is available for some autoimmune conditions, such as systemic sclerosis, and some outcomes, such as gestational diabetes mellitus (GDM)(^3^). To the best of our knowledge, no previous studies have been able to comprehensively explore the relation of most common autoimmune conditions with a wide range of pregnancy outcomes.

Mendelian randomization (MR) leverages genetic variants to test the causal effect of potential risk factors on health outcomes(^6^). Since genetic variants are fixed at conception and randomly assorted at meiosis, MR mitigates bias due to reverse causation and common confounders, such as socioeconomic position(^7^). Consistent with observational evidence, MR studies have suggested that liability to rheumatoid arthritis and systemic lupus erythematosus may cause increased risks of PTB and preeclampsia(^8–10^), but these studies have not considered a range of autoimmune conditions with a wide range of pregnancy outcomes, accounted for potential biases due to fetal genetics, or explored the role of the highly pleiotropic human leukocyte antigen (HLA) genetic region.

Our aim was to determine potential causal effects of liability to autoimmune conditions on pregnancy outcomes using MR. These analyses should be interpreted qualitatively, as previous MR studies with binary condition exposures have done. A precise, statistically robust result suggesting higher odds provides evidence that the condition causes an increased risk of the outcome, but the magnitude of the effect here is not clinically interpretable.

## Methods

We followed the Strengthening the Reporting of Observational Studies in Epidemiology using Mendelian Randomization (STROBE-MR) reporting guidelines(^11^).

We used two-sample MR(^6^) to explore the causal effects of liability to 10 autoimmune conditions on nine prespecified primary and seven secondary pregnancy outcomes. Genetic association data for autoimmune conditions were obtained from previous genome-wide association studies (GWAS) (n cases = 5,201 – 47,429, n controls = 9,066 – 501,638) while genetic data for pregnancy outcomes were obtained from the MR-PREG collaboration (n =77,285 – 934,566)(^12^). **Figure 1** outlines the methodological steps, with details of each step provided below.

**Figure 1.**
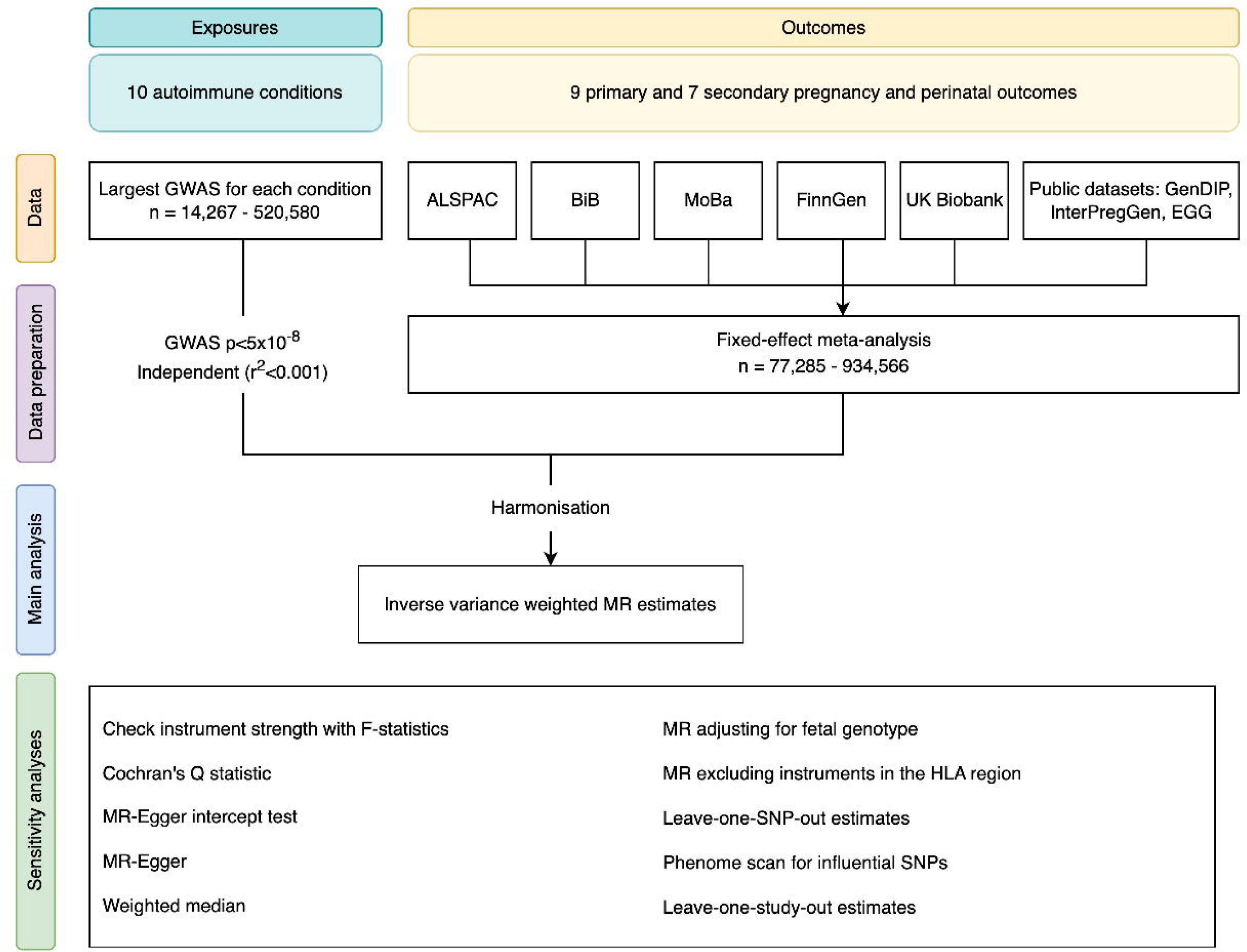
Outline of study design illustrating data sources, data preparation and analyses. GWAS = genome-wide association study, ALSPAC = Avon Longitudinal Study of Parents and Children, BiB = Born in Bradford, MoBa = the Norwegian Mother, Father and Child Cohort Study, GenDIP = GENetics of Diabetes In Pregnancy, InterPregGen = International Pregnancy Genetics study, EGG = Early Growth Genetics consortium, MR = Mendelian randomization, HLA = human leukocyte antigen, SNP = single nucleotide polymorphism.

We selected autoimmune conditions on the basis that they may be diagnosed in women of reproductive age(^1^) and that there was at least one published GWAS of the condition with at least 5,000 cases to ensure our MR analyses were well-powered (**Figure S1**). We identified the largest GWAS for each condition on GWAS Catalogue(^13^) and OpenGWAS(^14^). Further details of these GWAS, including references, can be found in **Table S1**. This identified 10 conditions meeting our criteria (**Figure S1**): ankylosing spondylitis, coeliac disease, Hashimoto’s thyroiditis, inflammatory bowel disease, multiple sclerosis, psoriasis, rheumatoid arthritis, systemic lupus erythematosus, systemic sclerosis, and type 1 diabetes.

### Instrument selection

For each GWAS, variants associated with the respective autoimmune condition (p-value < 5x10^−8^) were clumped to select independent genetic instruments (linkage disequilibrium threshold R^2^ < 0.001 using the 1000 genomes European super population reference panel). For systemic sclerosis full summary statistics were not available so the reported independent variants were selected, identified in the original GWAS by stepwise joint conditional analysis in 1.5Mb-windows around the lead single nucleotide polymorphism (SNP).

We explored potential effects on nine primary pregnancy outcomes, selected based on clinical importance and previously reported associations(^3–5,15^) (**Table S2**): miscarriage, stillbirth, HDP, GDM, PTB, small-for-gestational-age (SGA), large-for-gestational-age (LGA), low Apgar score at 5 minutes, and NICU admission. Seven secondary outcomes were also explored, including sub-categories of primary outcomes (e.g. preeclampsia and gestational hypertension as subtypes of HDP) and underlying continuous traits to provide more power and help interpretation of findings (e.g. birthweight). Outcome summary GWAS association data were obtained from the MR-PREG collaboration which integrates data from five cohort studies and four publicly available GWAS, including up to 934,566 pregnancies(^12^). Outcomes, definitions and sample sizes can be found in **Tables S2A-B**, and cohort descriptions in **Supplementary material**.

### Statistical analysis

Two-sample MR was conducted using the TwoSampleMR R package(^14^). All analyses were conducted in R (4.5.0). All code and a pre-specified analysis plan (dated 12/02/2024) are available at https://github.com/eaiton/autoimmune-pregnancy.

For our main analyses, we used the random-effects inverse-variance weighted (IVW) estimator. Effect estimates reflect the odds ratio (binary outcomes) or differences in mean standard deviation units (continuous outcomes) per doubling in genetically predicted log odds of the autoimmune condition. We scaled effect estimates and standard errors by multiplying these by log_e_2 before estimating 95% confidence intervals(^16^). Since our outcomes are rare, we assume odds and risks of outcomes are interchangeable. We identify plausible causal effects for follow up with sensitivity analyses with either (i) statistical support at p<0.05 or (ii) effect estimates larger than a 5% relative difference (i.e. odds ratio per doubling of genetic liability of >1.05 for higher risk and <0.95 for lowering risk) and with statistical support at p<0.10. We report whether effects passed multiple testing correction using the Benjami-Hochberg false discovery rate (alpha=0.05)(^17^), though our tests are not independent due to overlap between autoimmune condition genetic risk loci(^18^) and correlations between pregnancy outcomes.

We undertook a series of additional and sensitivity analyses to explore MR assumptions and potential bias in our main analyses. We used the pseudo F-statistic for each exposure as a measure of instrument strength (**Supplementary material**). We explored between SNP heterogeneity, which could occur through violation of any of the key MR assumptions, using Cochran’s Q-statistics. We explored unbalanced horizontal pleiotropy using two pleiotropy-robust MR methods: MR-Egger and weighed median analyses. Fetal genetic variants could also generate an independent path from the maternal genetic instruments to pregnancy outcomes. We explored this by comparing the main unadjusted results to results with adjustment for fetal genotype. First, we generated a GWAS estimating the effect of fetal genotype at each SNP on all available outcomes, using offspring genotypes available in the Avon Longitudinal Study of Parents and Children (ALSPAC), Born in Bradford (BiB) and the Norwegian Mother, Father and Child Cohort Study (MoBa). We then generated SNP-outcome associations adjusted for fetal genotype by combining this fetal genotype GWAS with the main maternal genotype GWAS in a weighted-linear model (WLM; see **Supplementary material**). We investigated whether specific studies in our outcome data were driving results through leave-one-study-out analyses.

We identified influential SNPs using leave-one-SNP-out analyses, and then performed a phenome-wide association scan on influential SNPs to identify any potential pleiotropic pathways, using OpenGWAS(^14^). In addition, we specifically investigated the potential role of the HLA genetic region on our results, given variants in this region are likely to relate to some of the autoimmune conditions we are exploring, and this region is highly pleiotropic and has an extensive linkage disequilibrium structure(^18^). We did this by excluding instruments within the HLA complex, defined as chr6:28,477,797-33,448,354 (genome reference build GRCh37).

## Results

Across the ten autoimmune conditions the mean pseudo-F-statistic ranged from 54 for systemic sclerosis to 286 for coeliac disease (**Table S3**). Main findings for primary outcomes are shown in **Figure 2** and for secondary outcomes in **Figures S2-3**. Results from all condition-outcome relationships and all sensitivities are provided in **Tables S4-S9**, for both 1-unit increase and doubling in log odds of autoimmune conditions.

**Figure 2.**
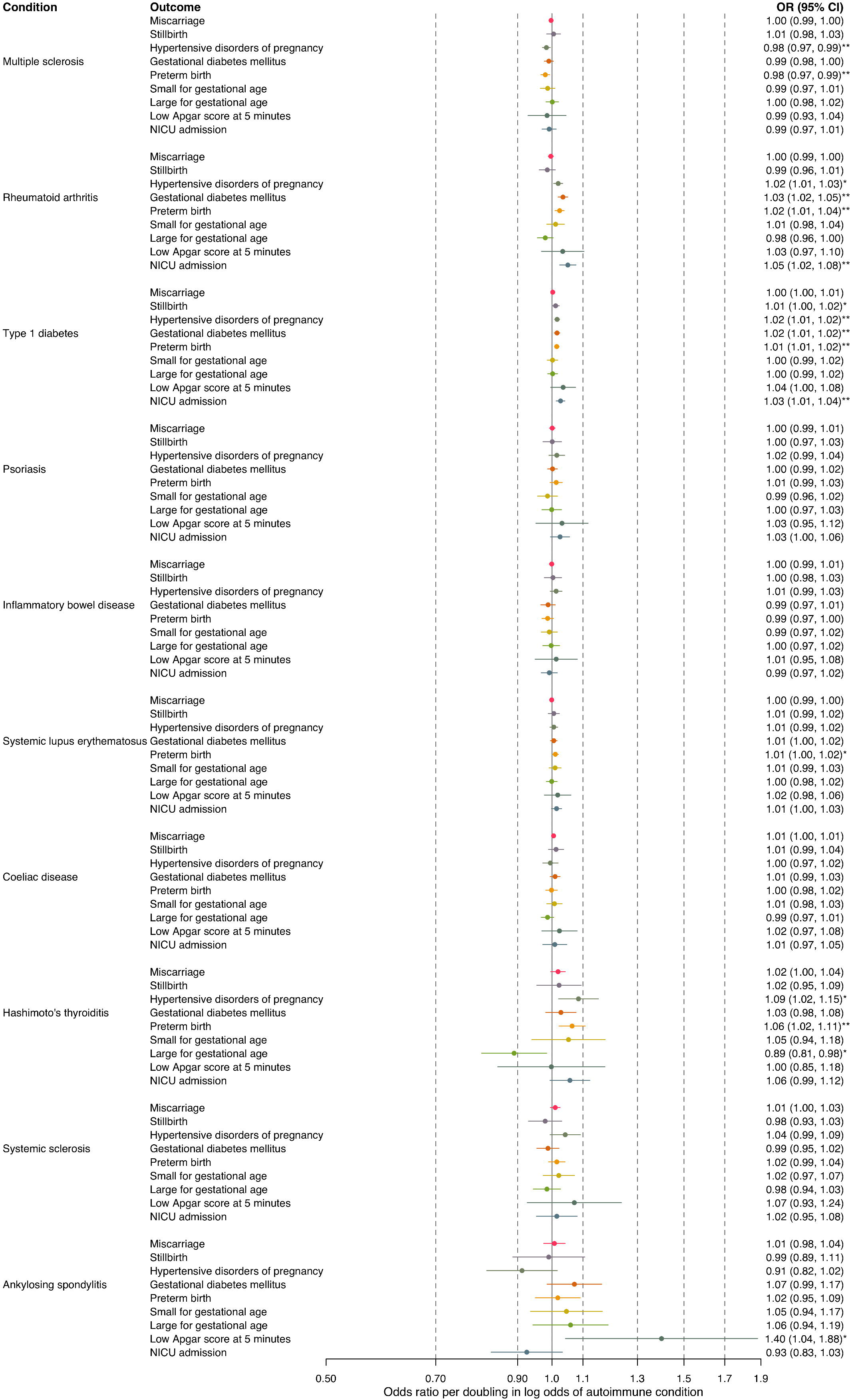
Mendelian randomization estimates for the effect of increased log odds of autoimmune conditions on primary pregnancy outcomes. Estimates reflect odds ratio of pregnancy outcome per doubling in log odds of autoimmune condition. NICU = neonatal intensive care unit. * indicates p<0.05, ** indicates p < 0.05 after correcting for multiple testing using false discovery rate.

We identified 18 effects for follow up (**Table 1**): 16 effects with statistical support at p<0.05, and two effects with a ≥5% change in odds and statistical support at p<0.1. Of these 18 effects, 14 were robust across sensitivity analyses and we focus on these here (**Table 1**, overall evidence ‘supportive’ since no sensitivity analyses found evidence of bias).

**Table 1.** Main analysis results for follow up and summary of sensitivity analyses. ^1^. Statistical support at * indicates p<0.05, ** indicates p < 0.05 after correcting for multiple testing using false discovery rate. ^2^Influential SNPs were identified in the leave-one-SNP-out analyses and a phenome scan was conducted to identify any influential SNPs which may be invalid due to plausible horizontal pleiotropic pathways. Cells stating ‘NA’ indicate where analyses could not be run since no SNPs were available in the outcome dataset, or outcome was not available. IVW = inverse variance weighted estimate.

| Autoimmune condition | Pregnancy outcome | MR direction | Statistical support<sup>1</sup> | Consistent MR methods | No bias from fetal genetics | No influential study | No influential SNPs<sup>2</sup> | Not driven by HLA | Overall evidence |
| --- | --- | --- | --- | --- | --- | --- | --- | --- | --- |
| Multiple sclerosis | HDP | - | * | ☐ | ☐ | ☐ | ☐ | ☐ | Inconclusive |
| Multiple sclerosis | PTB | - | ** | ☐ | ☐ | ☐ | ☐ | ☐ | Inconclusive |
| Rheumatoid arthritis | HDP | + | * | ☐ | ☐ | ☐ | ☐ | ☐ | Supportive |
| Rheumatoid arthritis | GDM | + | ** | ☐ | ☐ | ☐ | ☐ | ☐ | Supportive |
| Rheumatoid arthritis | PTB | + | ** | ☐ | ☐ | ☐ | ☐ | ☐ | Inconclusive |
| Rheumatoid arthritis | NICU admission | + | ** | ☐ | ☐ | ? | ☐ | ☐ | Supportive |
| Type 1 diabetes | Stillbirth | + | * | ☐ | NA | ? | ☐ | ☐ | Supportive |
| Type 1 diabetes | HDP | + | ** | ☐ | ☐ | ? | ☐ | ☐ | Supportive |
| Type 1 diabetes | GDM | + | ** | ☐ | ☐ | ☐ | ☐ | ? | Supportive |
| Type 1 diabetes | PTB | + | ** | ☐ | ☐ | ☐ | ☐ | ☐ | Inconclusive |
| Type 1 diabetes | NICU admission | + | ** | ☐ | ☐ | ☐ | ☐ | ☐ | Supportive |
| Systemic lupus erythematosus | PTB | + | * | ☐ | ☐ | ☐ | ☐ | ☐ | Supportive |
| Hashimoto's thyroiditis | LGA | - | * | ☐ | ☐ | ☐ | ? | NA | Supportive |
| Hashimoto's thyroiditis | HDP | + | * | ☐ | ☐ | ☐ | ☐ | ☐ | Supportive |
| Hashimoto's thyroiditis | PTB | + | ** | ☐ | ☐ | ☐ | ☐ | ☐ | Supportive |
| Hashimoto's thyroiditis | NICU admission | + |  | ☐ | ☐ | ☐ | ? | NA | Supportive |
| Ankylosing spondylitis | HDP | - |  | ☐ | ☐ | ☐ | ☐ | ☐ | Supportive |
| Ankylosing spondylitis | Low Apgar score | + | * | ? | ☐ | ? | ☐ | ☐ | Supportive |

**Table criteria:**
| Key | ☐ | ? | ☐ |
| --- | --- | --- | --- |
| Type of evidence | Supportive | Uncertain | Not supportive |
| **Consistency across MR methods** | Effect estimates from MR-Egger and weighted median in the same direction and of similar magnitude to IVW estimate | Effect estimates from MR-Egger and weighted median too imprecise to confidently determine whether the result is supportive or not | Effect estimates from MR-Egger and weighted median in the opposite direction or different magnitude to IVW estimate |
| **No bias by fetal genetics** | Maternal effect estimates accounting for fetal genotype in the same direction and of similar magnitude to IVW estimate | Maternal effect estimates accounting for fetal genotype too imprecise to confidently determine whether the result is supportive or not | Maternal effect estimate accounting for fetal genotype markedly attenuates or in the opposite direction to IVW estimate, **and** strong evidence of direct fetal genetic effects in the same direction as maternal effects |
| **No influential study** | Effect estimates when removing each study at a time in the same direction and of similar magnitude to the combined IVW estimate | Effect estimates when removing a study too imprecise to confidently determine whether the result is supportive or not | Effect estimate when removing a study markedly attenuates or in the opposite direction to combined IVW estimate |
| **No influential SNPs** | Effect estimates without each SNP all in the same direction and of similar magnitude to IVW estimate, **or** influential SNP identified and phenome scan for this influential SNP identified no plausible horizontal pleiotropic path | Effect estimates without each SNP too imprecise to confidently determine whether the result is supportive or not | Effect estimate without one SNP markedly attenuates or in the opposite direction to IVW estimate **and** phenome scan for this influential SNP identified plausible horizontal pleiotropic path |
| **Not driven by HLA** | Effect estimate without HLA variants are in the same direction and of similar magnitude to IVW estimate | Effect estimate without HLA variants too imprecise to confidently determine whether the result is supportive or not, including partial attenuation | Effect estimate without HLA variants markedly attenuates or in the opposite direction to IVW estimate |

We observed a lower risk of some pregnancy outcomes related to higher liability to both Hashimoto’s thyroiditis and ankylosing spondylitis. Higher liability to Hashimoto’s thyroiditis reduced risks of LGA (OR=0·89 (95%CI: 0·81, 0·98) per doubling in log odds or OR=0·85 (0·73, 0·98) per 1-unit increase in log odds). Higher ankylosing spondylitis liability reduced risks of HDP (0.91 per doubling in log odds (0·82, 1·02)).

We observed a higher risk of at least one adverse pregnancy outcome with higher liability to rheumatoid arthritis, type 1 diabetes, systemic lupus erythematosus, Hashimoto’s thyroiditis and ankylosing spondylitis. For example, higher liability to rheumatoid arthritis robustly increased risks of three outcomes, such as GDM (1·03 per doubling in log odds (1·02, 1·05)). Similarly, type 1 diabetes liability increased risks of four outcomes, including NICU admission (1·03 (1·01, 1·04)). A doubling in log odds of systemic lupus erythematosus increased risks of PTB only (1·01 (1·00, 1·02)), while ankylosing spondylitis liability increased risks of low Apgar score at 5 minutes (1·40, (1·04, 1·88)) only. Higher Hashimoto’s thyroiditis liability increased risks of three outcomes, such as PTB (1·06 (1·02, 1·11)).

### Secondary outcomes

Conditions that affected odds of combined HDP also affected preeclampsia and gestational hypertension separately (rheumatoid arthritis, type 1 diabetes, and Hashimoto’s thyroiditis, ankylosing spondylitis) (**Figures S2-3**).

Similarly, where higher liability to conditions increased risk of PTB and/or decreased risk of LGA as primary outcomes, these findings were supported by findings for underlying related secondary outcomes. Specifically, higher liability to systemic lupus erythematosus and Hashimoto’s thyroiditis both decreased mean gestational age at birth and birthweight (**Figure S3**). Estimated effects of Hashimoto’s thyroiditis liability on high birthweight and low birthweight were directionally concordant with higher liability decreasing LGA risk, though 95% confidence intervals were imprecise for these outcomes (**Figure S2**).

For conditions that increased the odds of PTB, we saw similar increases in risks of spontaneous PTB as a secondary outcome (**Figure S2**). Point estimates for increased liability were very similar for all and spontaneous PTB for both increased systemic lupus erythematosus liability (all PTB: 1·01 (1·00, 1·02); spontaneous PTB: 1·01 (1·00, 1·02)) and Hashimoto’s thyroiditis liability (all PTB: 1·06 (1·02, 1·12); spontaneous PTB: 1·06 (1·01, 1·12).

### Sensitivity analyses

**Table 1** summarises support from sensitivity analyses for the 18 causal condition-primary outcome relationships we identified for follow-up based on statistical significance and/or effect magnitude.

There was evidence of substantial between-SNP heterogeneity for 12 of the 18 condition-outcome relationships (p<0.5) (**Table S4**). However, MR-Egger and weighted median effect estimates were largely compatible with main IVW results (**Figure S4**). MR-Egger intercepts suggested potential unbalanced pleiotropy for two relationships (p<0.05): type 1 diabetes liability on stillbirth (p=0.032, intercept=-2.06x10^-3^) and on PTB (p=0.034, intercept=-1.82x10^-2^), but MR-Egger effect estimates were compatible with IVW estimates for both outcomes.

After adjustment for fetal genotypes in ALSPAC, BiB and MoBa, our effect estimates were consistent with the main unadjusted results, though because of the smaller sample size were less precise (**Figure S5**). Similarly, leave-one-study-out analyses were compatible with main meta-analysis results (**Figures S6-S12**), and showed anticipated imprecision when the largest cohort contributing to each outcome was left out (for example, UK Biobank for stillbirth, MoBa for low Apgar score at five minutes and NICU admission).

Of the 18 condition - primary outcome effects selected for follow up, four were influenced by one SNP which when removed markedly attenuated or changed the direction of the effect estimate, and two were uncertain (**Figures S13-S30**; **Table 2**). All four effects involved one of three SNPs, all located in the HLA region. The phenome scan suggested that these SNPs affected height or body mass index in the correct direction for horizontal pleiotropy to have plausibly biased these condition-outcome relationships (**Table 2; Figures S31-33**; for full results see **ST13**).

**Table 2.**
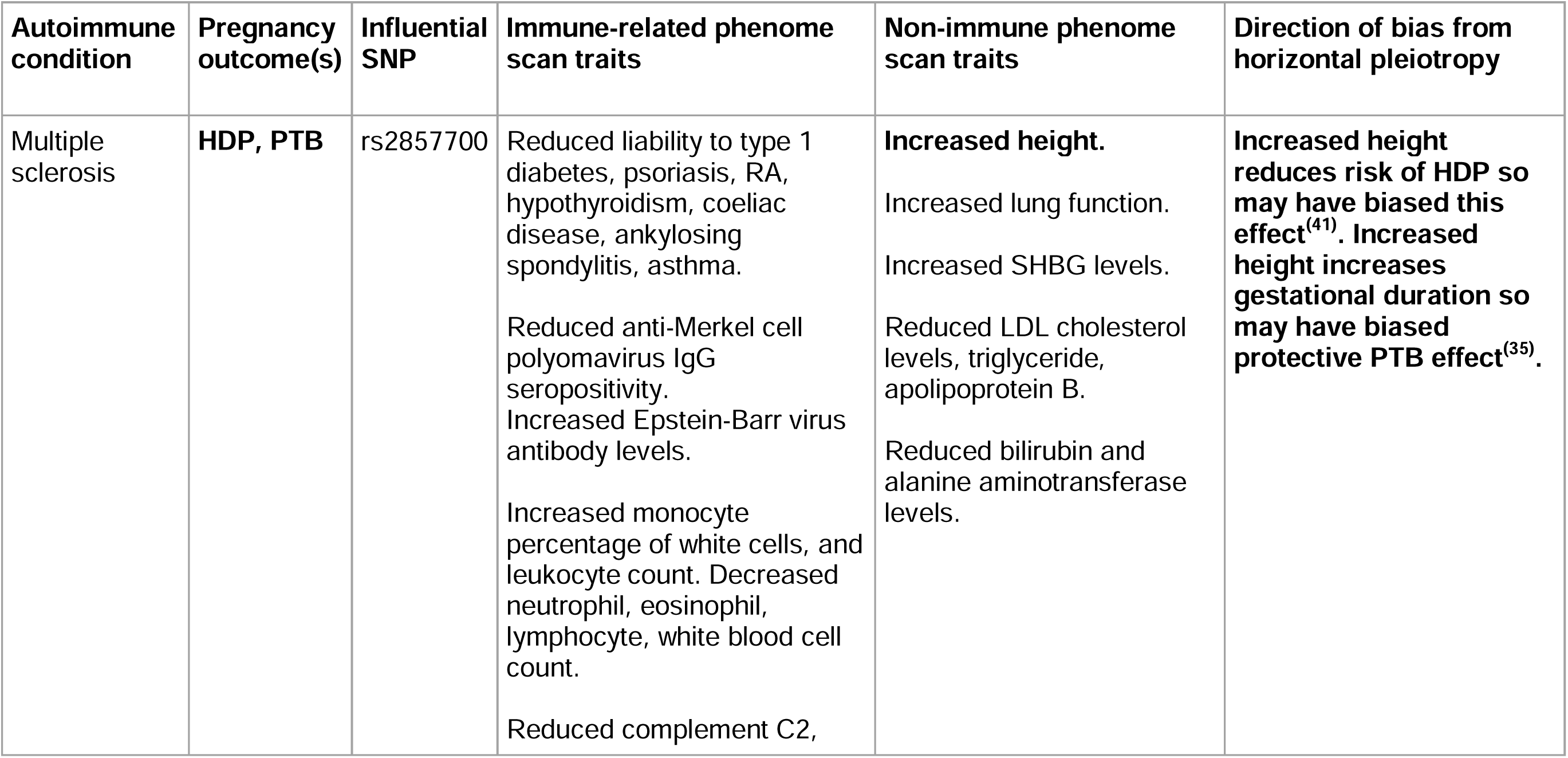
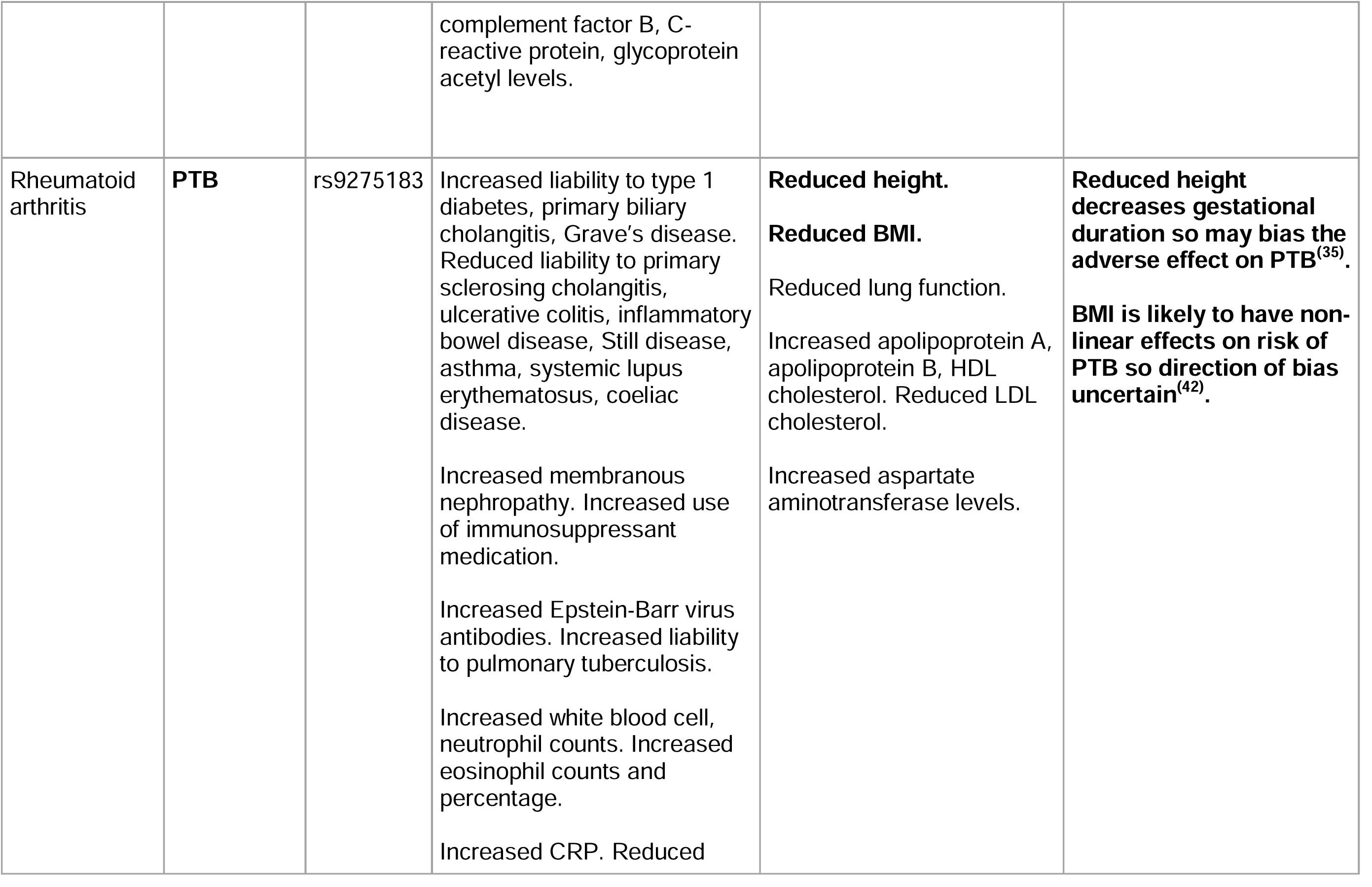
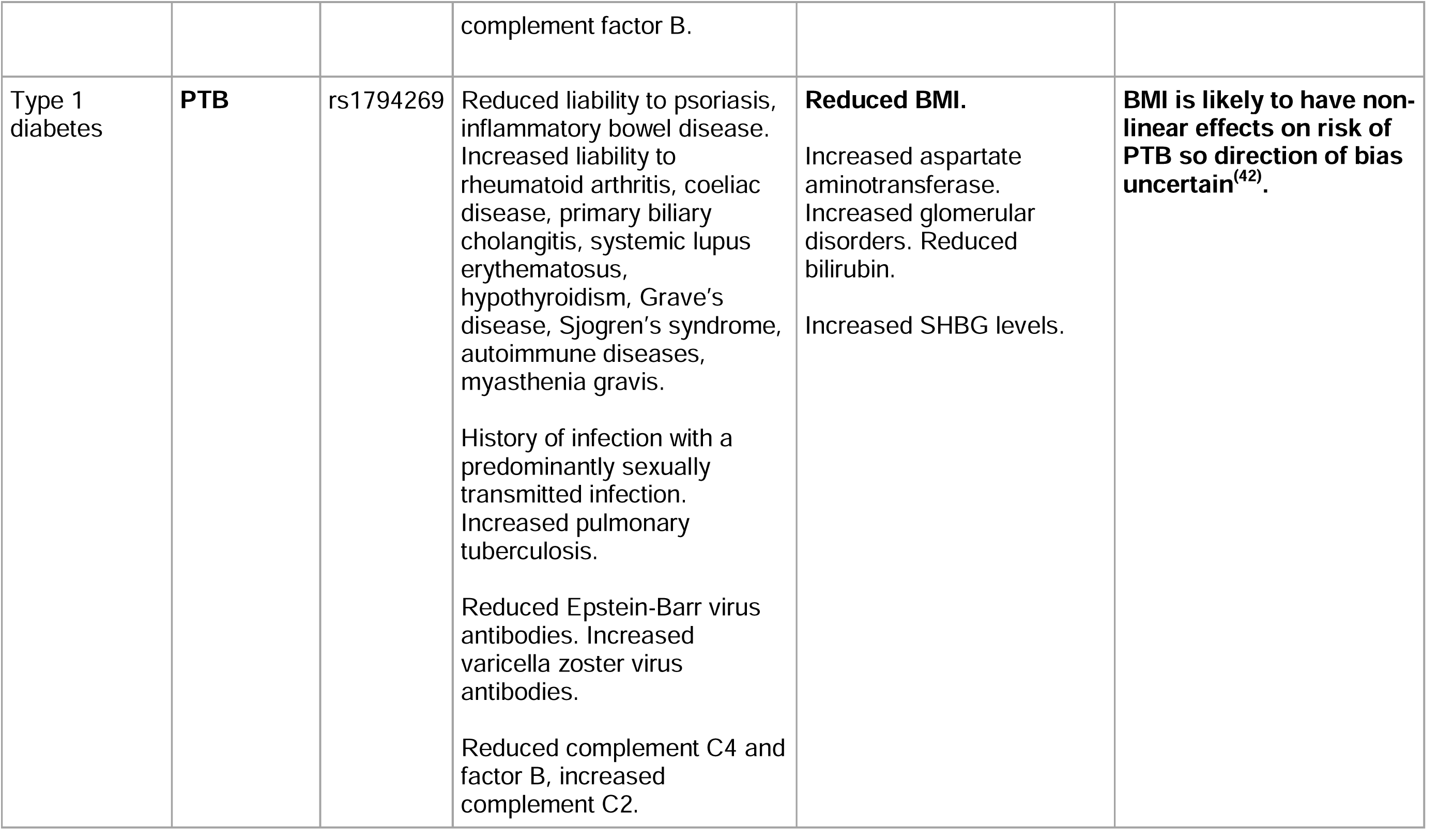
Influential SNPs driving the effects of autoimmune condition liability on pregnancy outcomes, and selected relevant associated traits from phenome-wide association scan. Bold text highlights where non-immune traits may lie on horizontal pleiotropic pathways since they are associated with the pregnancy outcomes also in bold, in the expected direction to have biased main results for one or more outcomes. Effects on other traits all given in terms of the effect allele increasing liability to the autoimmune condition exposure of interest. Traits selected based on relevance, removing duplicates, the exposure and the outcome(s) of interest and medications relevant to these. SHBG = sex hormone-binding globulin, LDL = low density lipoprotein, HDL = high density lipoprotein, CRP = C-reactive protein.

Across autoimmune conditions, 0-11 SNPs were in the HLA region, comprising from none (systemic sclerosis) up to 33% (coeliac disease, n=5/15) of the instruments. Excluding these SNPs reduced precision but largely produced effect estimates of similar magnitude and direction to the main analysis (**Figure S4**).

Of the 18 condition-primary outcome relationships followed up, the 14 causal effects which were robust across sensitivity analyses are summarised in **Table 3**.

**Table 3.**
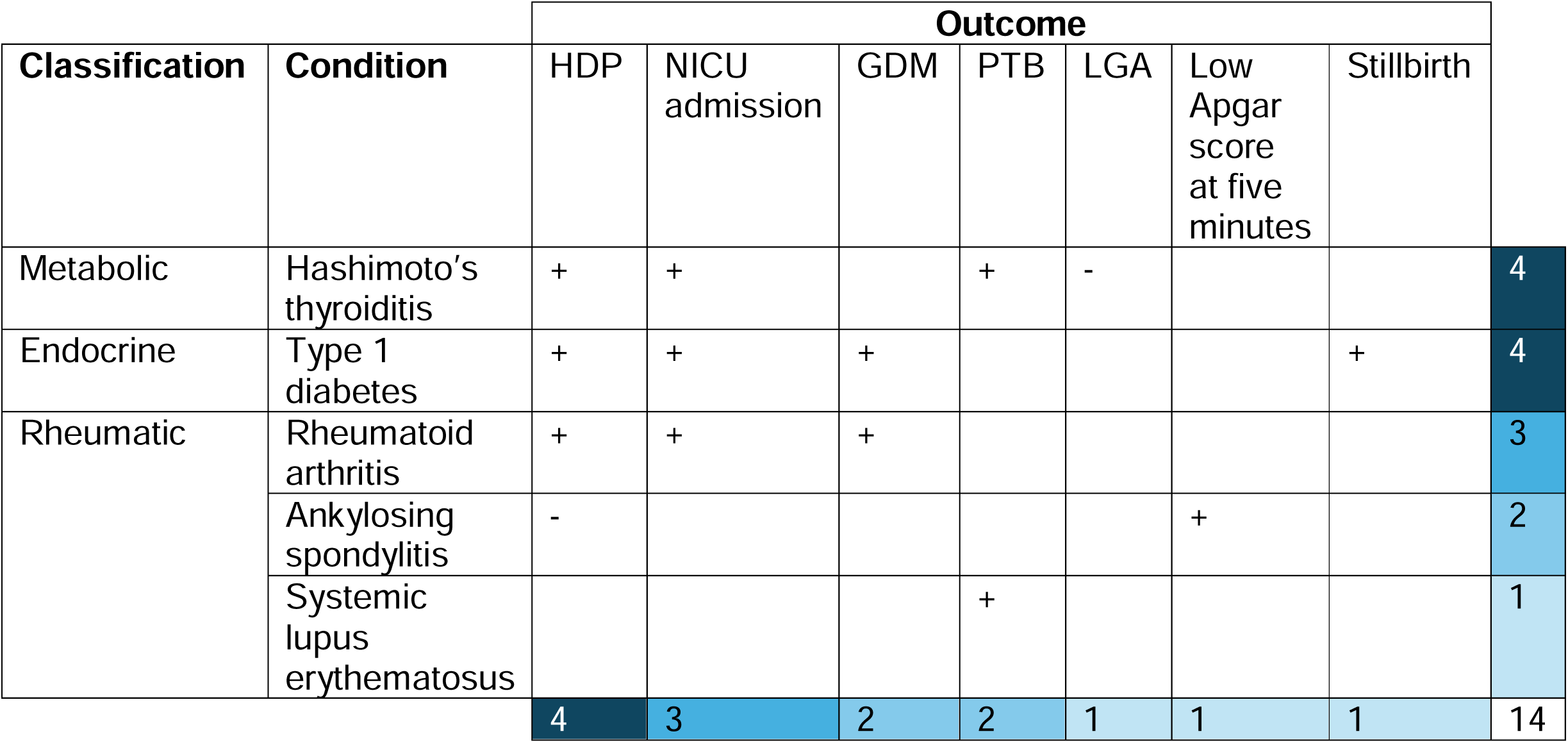
Summary of robust effects across sensitivity analyses. Causal effects where one or more sensitivity analyses was uncertain or could not be performed have been included, and effects which may be driven by HLA are also included. - indicates that higher liability has a protective effect, + indicates that higher liability has an adverse effect on risk of outcome. Numbers are counts of robust effects for each condition (rows) and each outcome (columns).

## Discussion

Previous conventional multivariable regression analyses have mostly focused on relationships between a limited number of autoimmune conditions and adverse pregnancy outcomes(^2–5^). By contrast we have explored potential causal effects of liability to a range of autoimmune conditions on numerous adverse pregnancy outcomes. These results show diverse associations across conditions and outcomes, with some evidence of genetic liability to conditions resulting in lower risk of some adverse outcomes and higher risk of other adverse outcomes (Hashimoto’s thyroiditis, ankylosing spondylitis), liability to other conditions only resulting in higher risk of adverse outcomes (rheumatoid arthritis, type 1 diabetes, systemic lupus erythematosus), and some conditions for which we found no evidence of robust effects on the outcomes we explored (multiple sclerosis, systemic sclerosis, coeliac disease, inflammatory bowel disease and psoriasis).

Our results are consistent with previous MR studies which estimated effects of systemic lupus erythematosus, rheumatoid arthritis and inflammatory bowel disease liability on selected pregnancy outcomes in FinnGen, one of our contributing cohorts(^8,9,19,20^).

Differences in the number of autoimmune conditions and pregnancy outcomes analysed, the effects estimated, and statistical power between our study and previous observational studies make direct comparisons difficult. Our causal estimates represent effects (odds ratios or difference in means) per doubling in the log odds of the lifetime genetic liability within the general population, so cannot be directly compared with observational studies considering the effects of having a diagnosed autoimmune condition. We therefore recommend interpretation of effects based on their direction rather than their magnitude, which has no direct clinical interpretation.

Positive observational associations of Hashimoto’s thyroiditis with HDP, NICU admission, and PTB; type 1 diabetes with HDP, NICU admission, and stillbirth; rheumatoid arthritis with HDP, NICU admission, and GDM; and systemic lupus erythematosus with PTB, are consistent with our evidence of causal effects for these(^3,21^). By contrast, our findings that liability to Hashimoto’s thyroiditis decreased risks of LGA(^22^), and that ankylosing spondylitis decreased risks of HDP(^23^) and increased risks of low Apgar score at five minutes(^24^), were not found in observational studies. The differences between our MR findings and observational studies may be attributable to differences in the exposures under investigation, differences in power due to small sample sizes of women diagnosed with rarer conditions, and/or to factors such as treatment effects or residual confounding driving the observational associations.

Increased liability to rheumatic autoimmune conditions may affect pregnancy outcomes through two key mechanisms: elevated inflammation and autoantibodies(^25^). Systemic inflammation could contribute to disordered placentation in preeclampsia and to insulin resistance in GDM(^26^) via endothelial dysfunction, since both complications are strongly associated with decreased maternal flow mediated dilation(^27^). Proinflammatory markers are also associated with fetal growth, for instance higher IL-6 concentrations are associated with lower offspring birth weight both in women with rheumatoid arthritis(^25^) and women without any chronic conditions(^28^). The excess risk of PTB in rheumatic conditions is likely mediated by complications, including preeclampsia and infection during pregnancy(^29^), suggesting these could contribute here to the effect of SLE liability on PTB. Neonatal effects on NICU admission and low Apgar score may in turn be attributable to PTB, or could reflect consequences of intrauterine infections which account for 25-40% of spontaneous PTB cases, particularly earlier PTB(^30^), and have a range of fetal consequences(^31^). Antiphospholipid autoantibodies, frequently present in women with systemic lupus erythematosus and rheumatoid arthritis(^32^), are involved in inflammatory placental damage contributing to risks of both HDP and PTB(^25^). These autoantibodies are also more prevalent in women who have experienced recurrent pregnancy losses(^33^), suggesting potential roles in the etiology of further adverse pregnancy outcomes.

Liability to Hashimoto’s thyroiditis may likewise affect adverse pregnancy outcomes through the development of antibodies to thyroid peroxidase, thyroglobulin, or thyroid-stimulating hormone receptors(^34^). These autoantibodies are present in 3-18% of pregnant women, and are associated with increased risk of PTB even in euthyroid pregnancies(^34^).

Adverse effects of type 1 diabetes during pregnancy could be mediated by insulin resistance and immune dysregulation. Hyperglycaemia is a risk factor for adverse pregnancy outcomes, even below clinical thresholds for treatment(^35^). Type 1 diabetes liability may also affect pregnancy through characteristic dysregulated peripheral immune responses which are antagonistic to the responses seen in a healthy pregnancy, including an increased Th1/Th2 ratio and increased IFN-γ expression from NK cells(^36^); these may affect neonatal infection risk and thereby NICU admission. Our finding that higher type 1 diabetes liability increased GDM risk could reflect pleiotropy, since variants influencing insulin production will affect both conditions. It may also reflect misclassification, since some women progressing towards type 1 diabetes or with latent autoimmune diabetes before pregnancy may be diagnosed with GDM due to the additional strain pregnancy places on beta cells, and then later be diagnosed with type 1 diabetes(^37^).

Our MR analysis provides a new line of biological evidence which should be less susceptible to conventional observational confounding(^7^) and reverse causation(^21^). The strengths of our approach include the systematic exploration of a range of autoimmune conditions and relevant pregnancy outcomes, our use of large-scale genetic cohort data to maximise power, and the consideration of fetal genetic effects and the HLA.

There were several limitations to this study. Since our exposure is genetic liability to a condition, rather than diagnosis, our results could reflect the effects of risk factors of an autoimmune condition rather than causal effects of disease status(^38^). This is particularly pertinent for conditions with lower prevalences(^16,38^), for instance systemic sclerosis and ankylosing spondylitis.

Our small effect sizes are likely the result of the fact that the conditions we investigate are rare and the resulting genetically predicted risk of each is relatively low(^1^). For some rarer outcomes we lack power, such as low Apgar score at five minutes. Even in large cohort studies of diagnosed patients, the estimated increased risk of adverse pregnancy outcomes from these conditions is often small to moderate (with the exception of type 1 diabetes(^21^)), so the causal effects of increased genetic liability may be too modest to be detectable without extremely large sample sizes, particularly for systemic sclerosis, inflammatory bowel disease, coeliac disease and psoriasis(^21^).

For eight autoimmune conditions of interest, no sufficiently large GWAS was publicly available. No exposure GWAS for autoimmune conditions were available stratified by sex, so we could not explore whether potential sex differences in instruments might bias our results. Except ankylosing spondylitis and type 1 diabetes, all conditions analysed here are more common in women which will mitigate this potential source of bias to some extent(^1^). Selection bias may have biased our estimates because some autoimmune conditions may reduce fertility(^21^). Since our study was restricted to women who have had at least one pregnancy, our results may therefore be subject to collider bias(^39^). We restricted to cohorts with majority white European ancestry, so further work is needed to assess whether these results are generalisable to non-white European ancestry groups. Our analyses may be biased by horizontal pleiotropy since autoimmune conditions are highly genetically correlated(^18^) and associated with health-related behaviours which may be tagged by large GWAS. We addressed this by assessing evidence for horizontal pleiotropy by exploring between-SNP heterogeneity, using pleiotropy-robust MR methods, excluding HLA variants, and by conducting a phenome scan for influential SNPs.

In conclusion, our study supports that increased genetic liability to Hashimoto’s thyroiditis, type 1 diabetes, rheumatoid arthritis, ankylosing spondylitis, and systemic lupus erythematosus increases the risks of adverse pregnancy outcomes, with effects particularly centred on HDP, GDM, PTB, and NICU admission.

Understanding whether reported associations between autoimmune conditions and adverse pregnancy outcomes are causal is important to inform clinical decision-making – if a condition itself increases risks, it suggests that discontinuation of treatments during pregnancy which manage symptoms may exacerbate the risks of adverse pregnancy outcomes. For instance, TNF-alpha inhibitors are now recommended throughout pregnancy to manage rheumatic conditions(^40^). Our findings underline that women with Hashimoto’s thyroiditis, type 1 diabetes and rheumatic conditions should have their conditions carefully managed during pregnancy to minimise complications.

## Data Sharing

All code and a pre-specified analysis plan (dated 12/02/2024) are available on GitHub at https://github.com/eaiton/autoimmune-pregnancy.

To protect participant confidentiality, supporting data cannot be made openly available. Researchers can apply for access to individual study executive committees.

The ALSPAC access policy describes the proposal process in detail including any costs associated with conducting research at ALSPAC, and may be updated from time to time (https://www.bristol.ac.uk/media-library/sites/alspac/documents/researchers/data-access/ALSPAC_Access_Policy.pdf). The ALSPAC study website contains details of all the data that is available through a fully searchable data dictionary and variable search tool (http://www.bristol.ac.uk/alspac/researchers/our-data/).

Data is available upon request from Born in Bradford (https://borninbradford.nhs.uk/research/how-to-access-data/).

Data from MoBa are available from the Norwegian Institute of Public Health after application to the MoBa Scientific Management Group (see its website https://www.fhi.no/en/ch/studies/moba/for-forskere-artikler/research-and-data-access/ for details).

Researchers can apply for access to the UK Biobank data via the Access Management System (AMS) (https://www.ukbiobank.ac.uk/enable-your-research/apply-for-access).

FinnGenn genetic summary statistics are freely available online (https://www.finngen.fi/en/access_results).

GWAS Catalogue (https://www.ebi.ac.uk/gwas/home) and IEU Open GWAS (https://gwas.mrcieu.ac.uk/) are publicly available resources.

## Author contributions

EA, MCB, and DAL conceived and designed the study. Study management and data collection for individual studies within the MR-PREG collaboration was conducted by DAL (ALSPAC; BiB), MCM (MoBa), and JW (BiB). EA, NM, GC, AGS, TB, QY, CC, and MCB undertook data curation for individual MR-PREG collaboration studies. QY and MCB ran the GWAS meta-analysis. TB and MCB generated the WLM models. EA wrote an initial analysis plan, with input from MCB and DAL. EA conducted the statistical analyses. EA wrote the first draft of the manuscript. All authors provided comments and critical revisions on drafts of the manuscript.

## Ethics

For all included studies, written informed consent was obtained from participants and approval was obtained from relevant ethics committees. Details on ethical approval for individual studies are provided in **Supplementary material**.

## Data Availability

All code and a pre-specified analysis plan (dated 12/02/2024) are available on GitHub at https://github.com/eaiton/autoimmune-pregnancy.
To protect participant confidentiality, supporting data cannot be made openly available. Researchers can apply for access to individual study executive committees.
The ALSPAC access policy describes the proposal process in detail including any costs associated with conducting research at ALSPAC, and may be updated from time to time (https://www.bristol.ac.uk/media-library/sites/alspac/documents/researchers/data-access/ALSPAC_Access_Policy.pdf). The ALSPAC study website contains details of all the data that is available through a fully searchable data dictionary and variable search tool (http://www.bristol.ac.uk/alspac/researchers/our-data/).
Data is available upon request from Born in Bradford (https://borninbradford.nhs.uk/research/how-to-access-data/).
Data from MoBa are available from the Norwegian Institute of Public Health after application to the MoBa Scientific Management Group (see its website https://www.fhi.no/en/ch/studies/moba/for-forskere-artikler/research-and-data-access/ for details).
Researchers can apply for access to the UK Biobank data via the Access Management System (AMS) (https://www.ukbiobank.ac.uk/enable-your-research/apply-for-access).
FinnGenn genetic summary statistics are freely available online (https://www.finngen.fi/en/access_results).
GWAS Catalogue (https://www.ebi.ac.uk/gwas/home) and IEU Open GWAS (https://gwas.mrcieu.ac.uk/) are publicly available resources.

## Supporting information

**S1. Supplementary material**. (PDF)

**S2. Supplementary tables**. (XLSX)

**S3. STROBE MR checklist**. (DOCX)

## References

1. Conrad N, Misra S, Verbakel JY, Verbeke G, Molenberghs G, Taylor PN, et al. Incidence, prevalence, and co-occurrence of autoimmune disorders over time and by age, sex, and socioeconomic status: a population-based cohort study of 22 million individuals in the UK. The Lancet. 2023 June 3;401(10391):1878–90.

2. Finkelsztejn A, Brooks J, Paschoal Jr F, Fragoso Y. What can we really tell women with multiple sclerosis regarding pregnancy? A systematic review and meta-analysis of the literature. BJOG: An International Journal of Obstetrics & Gynaecology. 2011;118(7):790–7.

3. Singh M, Wambua S, Lee SI, Okoth K, Wang Z, Fayaz FFA, et al. Autoimmune diseases and adverse pregnancy outcomes: an umbrella review. BMC Med. 2024 Dec;22(1):1–19.

4. Su PY, Huang K, Hao JH, Xu YQ, Yan SQ, Li T, et al. Maternal thyroid function in the first twenty weeks of pregnancy and subsequent fetal and infant development: a prospective population-based cohort study in China. J Clin Endocrinol Metab. 2011 Oct;96(10):3234–41.

5. Leung KK, Tandon P, Govardhanam V, Maxwell C, Huang V. The Risk of Adverse Neonatal Outcomes With Maternal Inflammatory Bowel Disease: A Systematic Review and Meta-analysis. Inflammatory Bowel Diseases. 2021 Apr 1;27(4):550–62.

6. Davies NM, Holmes MV, Smith GD. Reading Mendelian randomisation studies: a guide, glossary, and checklist for clinicians. BMJ. 2018 July 12;362:k601.

7. Smith GD, Lawlor DA, Harbord R, Timpson N, Day I, Ebrahim S. Clustered environments and randomized genes: a fundamental distinction between conventional and genetic epidemiology. PLoS Med. 2007 Dec;4(12):e352.

8. Yan P, Yao J, Ke B, Fang X. Mendelian randomization reveals systemic lupus erythematosus and rheumatoid arthritis and risk of adverse pregnancy outcomes. European Journal of Obstetrics & Gynecology and Reproductive Biology. 2024 Feb;293:78–83.

9. Chang T, Zhao Z, Liu X, Zhang X, Zhang Y, Liu X, et al. Rheumatoid arthritis and adverse pregnancy outcomes: a bidirectional two-sample mendelian randomization study. BMC Pregnancy Childbirth. 2024 July 31;24(1):517.

10. Zhang D, Hu Y, Guo W, Song Y, Yang L, Yang S, et al. Mendelian randomization study reveals a causal relationship between rheumatoid arthritis and risk for pre-eclampsia. Front Immunol. 2022;13:1080980.

11. Skrivankova VW, Richmond RC, Woolf BAR, Yarmolinsky J, Davies NM, Swanson SA, et al. Strengthening the Reporting of Observational Studies in Epidemiology Using Mendelian Randomization: The STROBE-MR Statement. JAMA. 2021 Oct 26;326(16):1614–21.

12. McBride N, Clayton GL, Soares ALG, Yang Q, Bond TA, Taylor A, et al. Cohort Profile: The Mendelian Randomization in Pregnancy (MR-PREG) collaboration - Improving evidence for prevention and treatment of adverse pregnancy and perinatal outcomes [Internet]. medRxiv; 2025 [cited 2025 Mar 24]. p. 2025.03.22.25324447. Available from: https://www.medrxiv.org/content/10.1101/2025.03.22.25324447v1

13. Cerezo M, Sollis E, Ji Y, Lewis E, Abid A, Bircan KO, et al. The NHGRI-EBI GWAS Catalog: standards for reusability, sustainability and diversity. Nucleic Acids Research. 2025 Jan 6;53(D1):D998–1005.

14. Hemani G, Zheng J, Elsworth B, Wade KH, Haberland V, Baird D, et al. The MR-Base platform supports systematic causal inference across the human phenome. Elife. 2018 May 30;7:e34408.

15. Sim BL, Daniel RS, Hong SS, Matar RH, Ganiel I, Nakanishi H, et al. Pregnancy Outcomes in Women With Rheumatoid Arthritis: A Systematic Review and Meta-analysis. J Clin Rheumatol. 2023 Jan 1;29(1):36–42.

16. Burgess S, Labrecque JA. Mendelian randomization with a binary exposure variable: interpretation and presentation of causal estimates. Eur J Epidemiol. 2018 Oct 1;33(10):947–52.

17. Benjamini Y, Hochberg Y. Controlling the False Discovery Rate: A Practical and Powerful Approach to Multiple Testing. Journal of the Royal Statistical Society: Series B (Methodological). 1995 Jan 1;57(1):289–300.

18. Lincoln MR, Connally N, Axisa PP, Gasperi C, Mitrovic M, van Heel D, et al. Genetic mapping across autoimmune diseases reveals shared associations and mechanisms. Nat Genet. 2024 May;56(5):838–45.

19. Zhang D, Hu Y, Guo W, Song Y, Yang L, Yang S, et al. Mendelian randomization study reveals a causal relationship between rheumatoid arthritis and risk for pre-eclampsia. Front Immunol. 2022 Dec 12;13:1080980.

20. Zhang X, Wu X, Chen L, He L. Autoimmune diseases and risk of gestational diabetes mellitus: a Mendelian randomization study. Acta Diabetol. 2024 Feb 1;61(2):161–8.

21. Kerola AM, Palomäki A, Laivuori H, Laitinen T, Färkkilä M, Eklund KK, et al. Patterns of reproductive health in inflammatory rheumatic diseases and other immune-mediated diseases: a nationwide registry study. Rheumatology. 2024 Mar 20;keae122.

22. Derakhshan A, Peeters RP, Taylor PN, Bliddal S, Carty DM, Meems M, et al. Association of maternal thyroid function with birth weight: a systematic review and individual-participant data meta-analysis. Lancet Diabetes Endocrinol. 2020 June;8(6):501–10.

23. Maguire S, O’Dwyer T, Mockler D, O’Shea F, Wilson F. Pregnancy in axial spondyloarthropathy: A systematic review & meta-analysis. Seminars in Arthritis and Rheumatism. 2020 Dec 1;50(6):1269–79.

24. Park EH, Lee JS, Kim YJ, Lee SM, Jun JK, Lee EB, et al. Pregnancy outcomes in Korean women with ankylosing spondylitis. Korean J Intern Med. 2019 Sept 26;36(3):721–30.

25. Andreoli L, Andersen J, Avcin T, Chambers CD, Fazzi EM, Marlow N, et al. The outcomes of children born to mothers with autoimmune rheumatic diseases. The Lancet Rheumatology. 2024 Aug 1;6(8):e573–86.

26. Shoelson SE, Lee J, Goldfine AB. Inflammation and insulin resistance. J Clin Invest. 2006 July 3;116(7):1793–801.

27. McElwain CJ, Tuboly E, McCarthy FP, McCarthy CM. Mechanisms of Endothelial Dysfunction in Pre-eclampsia and Gestational Diabetes Mellitus: Windows Into Future Cardiometabolic Health? Front Endocrinol (Lausanne). 2020 Sept 11;11:655.

28. Francis EC, Li M, Hinkle SN, Chen J, Wu J, Zhu Y, et al. Maternal Proinflammatory Adipokines Throughout Pregnancy and Neonatal Size and Body Composition: A Prospective Study. Curr Dev Nutr. 2021 Oct;5(10):nzab113.

29. Bandoli G, Singh N, Strouse J, Baer RJ, Donovan BM, Feuer SK, et al. Mediation of Adverse Pregnancy Outcomes in Autoimmune Conditions by Pregnancy Complications: A Mediation Analysis of Autoimmune Conditions and Adverse Pregnancy Outcomes. Arthritis Care Res (Hoboken). 2020 Feb;72(2):256–64.

30. Goldenberg RL, Culhane JF, Iams JD, Romero R. Epidemiology and causes of preterm birth. The Lancet. 2008 Jan 5;371(9606):75–84.

31. Megli CJ, Coyne CB. Infections at the maternal–fetal interface: an overview of pathogenesis and defence. Nat Rev Microbiol. 2022 Feb;20(2):67–82.

32. El Hasbani G, Viola M, Sciascia S, Taher AT, Uthman I. Antiphospholipid Antibodies in Inflammatory and Autoimmune Rheumatic and Musculoskeletal Diseases Beyond Lupus: A Systematic Review of the Available Evidence. Rheumatol Ther. 2021 Jan 9;8(1):81–94.

33. Carp HJA, Meroni PL, Shoenfeld Y. Autoantibodies as predictors of pregnancy complications. Rheumatology. 2008 June 1;47(suppl_3):iii6–8.

34. De Leo S, Pearce EN. Autoimmune thyroid disease during pregnancy. Lancet Diabetes Endocrinol. 2018 July;6(7):575–86.

35. Chen J, Bacelis J, Sole-Navais P, Srivastava A, Juodakis J, Rouse A, et al. Dissecting maternal and fetal genetic effects underlying the associations between maternal phenotypes, birth outcomes, and adult phenotypes: A mendelian-randomization and haplotype-based genetic score analysis in 10,734 mother–infant pairs. PLOS Medicine. 2020 Aug 25;17(8):e1003305.

36. Groen B, Links TP, van den Berg PP, de Vos P, Faas MM. The role of autoimmunity in women with type 1 diabetes and adverse pregnancy outcome: A missing link. Immunobiology. 2019 Mar 1;224(2):334–8.

37. Auvinen AM, Luiro K, Jokelainen J, Järvelä I, Knip M, Auvinen J, et al. Type 1 and type 2 diabetes after gestational diabetes: a 23 year cohort study. Diabetologia. 2020;63(10):2123–8.

38. Chen S, Liang Y, Mo JMY, Li QHY, He B, Luo S, et al. Challenges in interpreting Mendelian randomization studies with a disease as the exposure: Using COVID-19 liability studies as an exemplar. Eur J Hum Genet. 2025 May;33(5):658–65.

39. Griffith GJ, Morris TT, Tudball MJ, Herbert A, Mancano G, Pike L, et al. Collider bias undermines our understanding of COVID-19 disease risk and severity. Nat Commun. 2020 Nov 12;11(1):5749.

40. Russell MD, Dey M, Flint J, Davie P, Allen A, Crossley A, et al. British Society for Rheumatology guideline on prescribing drugs in pregnancy and breastfeeding: immunomodulatory anti-rheumatic drugs and corticosteroids. Rheumatology. 2023 Apr 1;62(4):e48–88.

41. Lee Y, Magnus P. Maternal and Paternal Height and the Risk of Preeclampsia. Hypertension. 2018 Apr;71(4):666–70.

42. Borges MC, Clayton GL, Freathy RM, Felix JF, Fernández-Sanlés A, Soares AG, et al. Integrating multiple lines of evidence to assess the effects of maternal BMI on pregnancy and perinatal outcomes. BMC Med. 2024 Jan 29;22(1):32.

