## Supplementary material for "Characterising the effects of genetic liability to autoimmune conditions on pregnancy outcomes using Mendelian Randomization"

#### Autoimmune condition selection

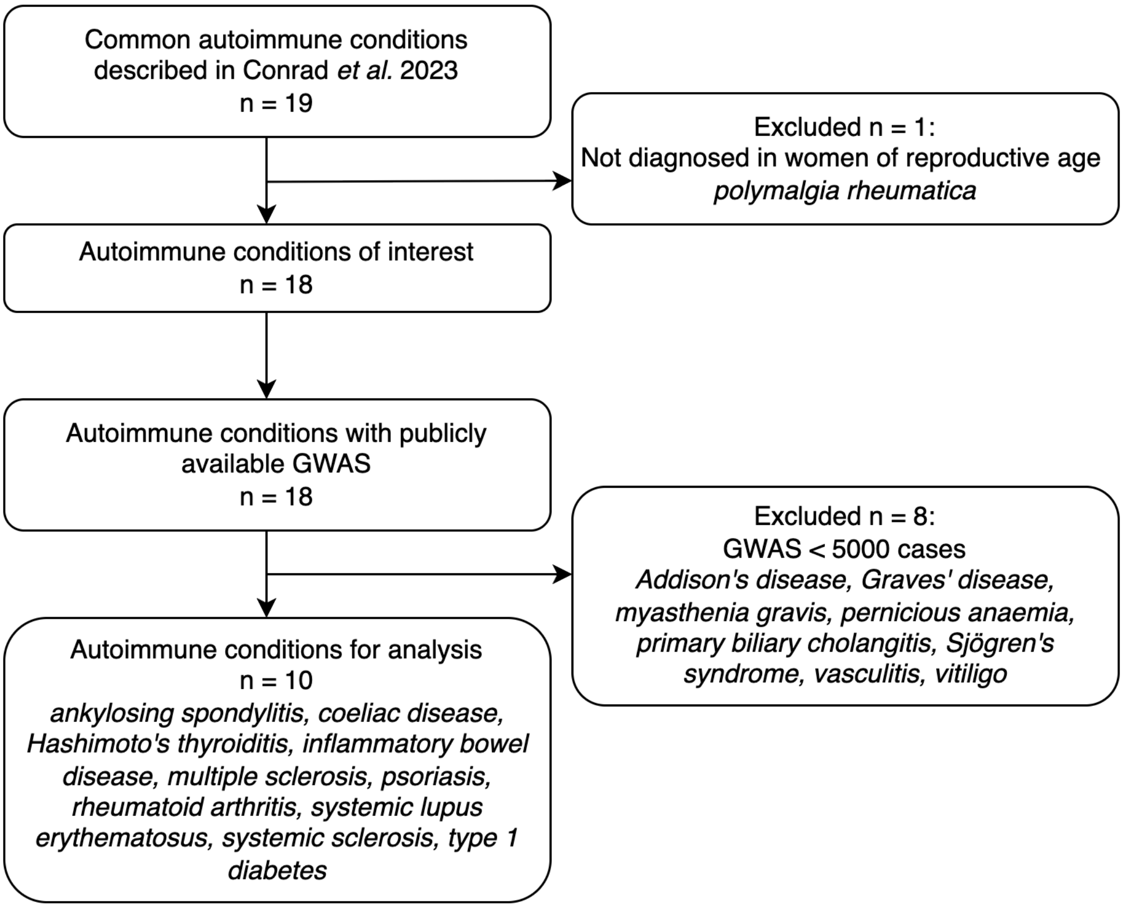

*Figure 1. Flow diagram explaining selection of autoimmune conditions for analysis. GWAS = genome-wide association study.*

#### Secondary outcomes

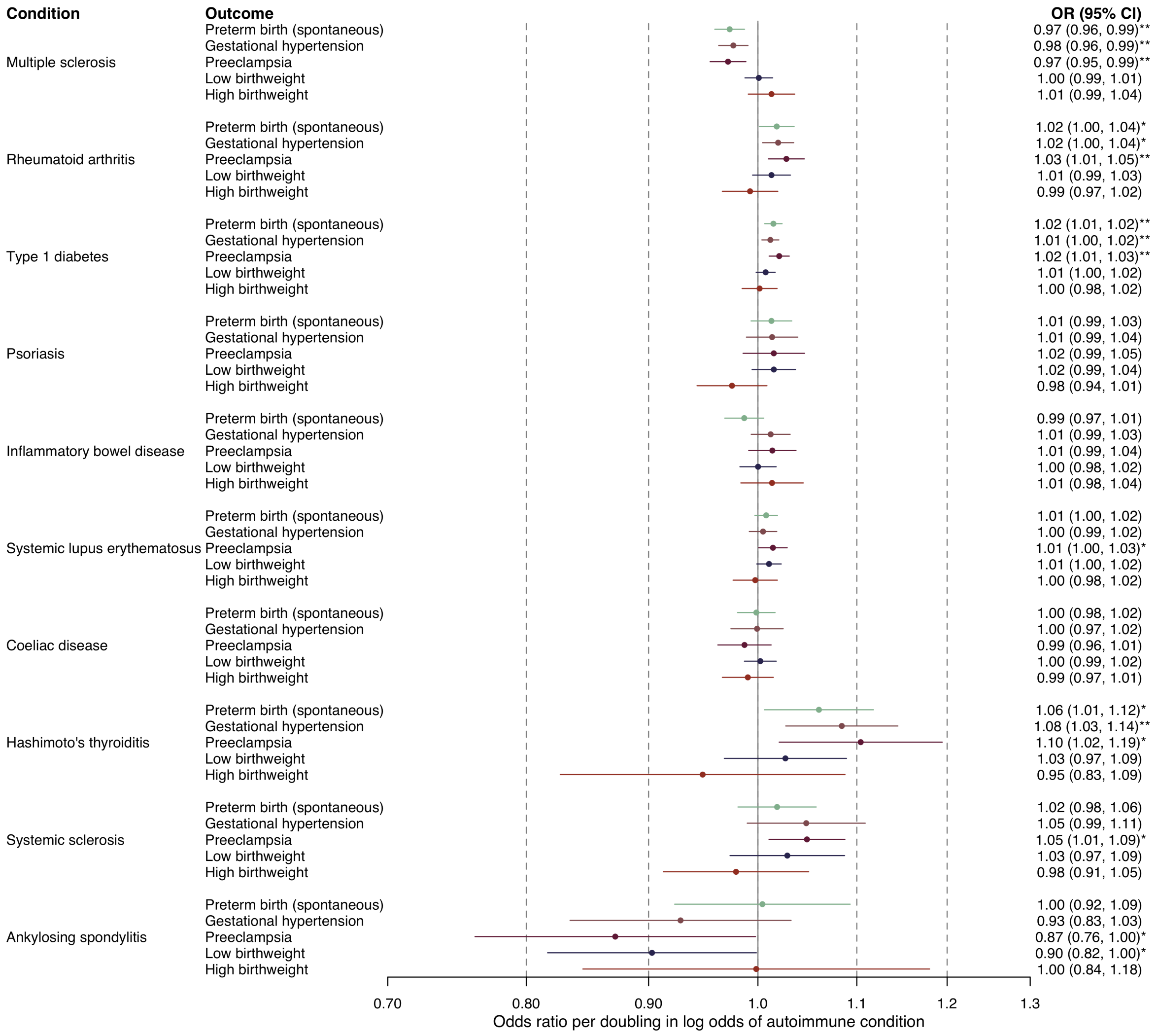

*Figure 2. Mendelian randomization estimates for the effect of log odds of autoimmune conditions on binary secondary pregnancy outcomes. Estimates reflect odds ratio of pregnancy outcome per doubling in log odds of autoimmune condition. * indicates p<0.05, ** indicates p < 0.05 after correcting for multiple testing using false discovery rate.*

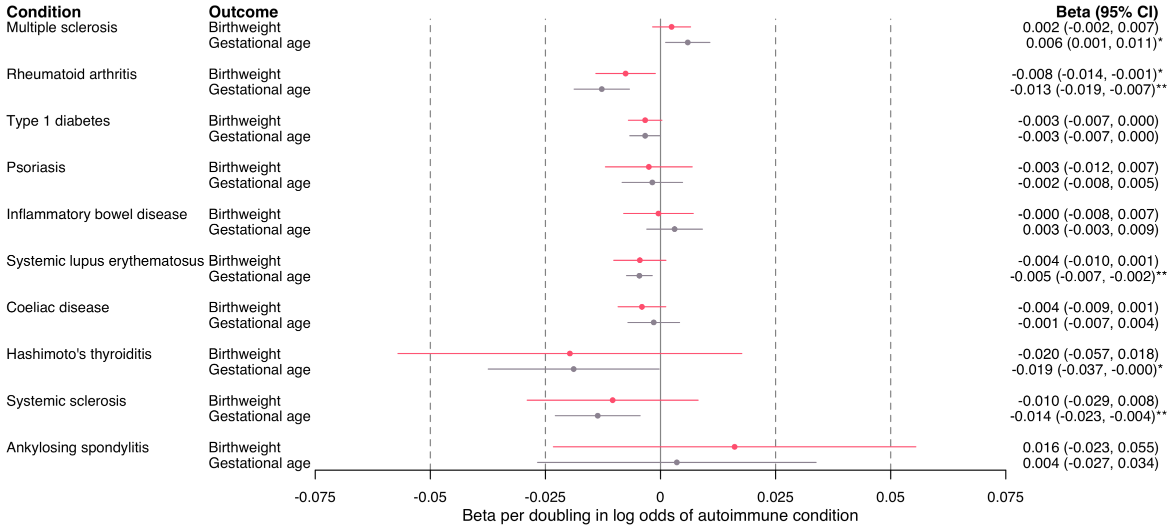

*Figure 3. Mendelian randomization estimates for the effect of log odds of autoimmune conditions on continuous secondary pregnancy outcomes. Estimates reflect mean difference in outcome (in standard deviation units) per doubling in log odds of autoimmune conditions. One SD birthweight is ~601.9 grams and one SD of gestational age is 1.8 weeks or 12.6 days. * indicates $p<0.05$, ** indicates $p < 0.05$ after correcting for multiple testing using false discovery rate.*

#### Pleiotropy-robust MR methods and excluding HLA variants

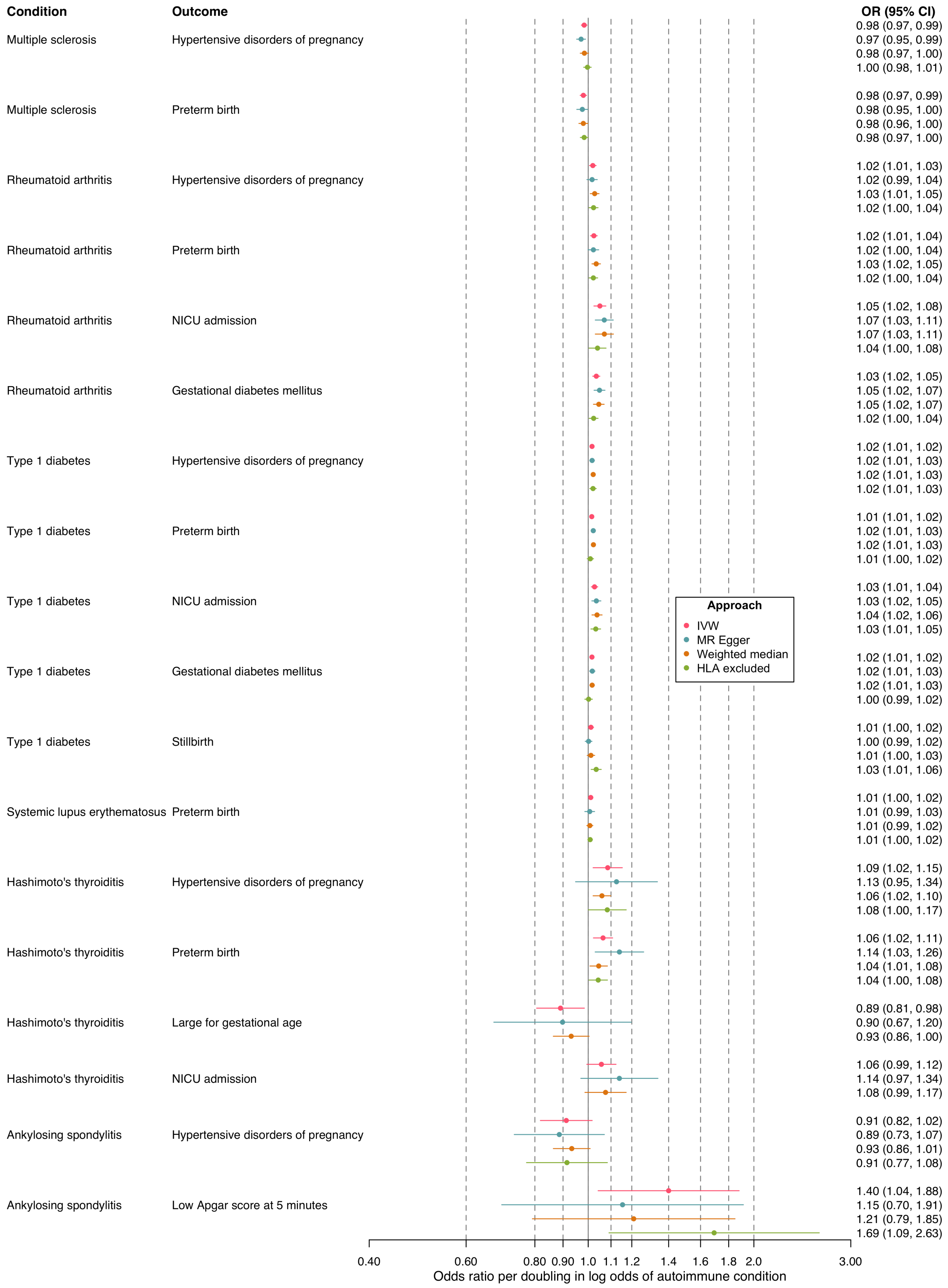

*Figure 4. Pleiotropy-robust methods and HLA excluded Mendelian randomization estimates for the effects of increased log odds of autoimmune conditions on pregnancy outcomes, for results selected for follow up. Estimates reflect odds ratio of pregnancy outcome per doubling in log odds of autoimmune condition. NICU = neonatal intensive care unit. IVW = inverse variance weighted. Effects excluding HLA could not be estimated for two effects of Hashimoto's thyroiditis liability since no HLA SNPs were available in these outcome datasets (n = 2 SNPs in HLA for Hashimoto's thyroiditis instruments).*

#### Fetal genetic effects

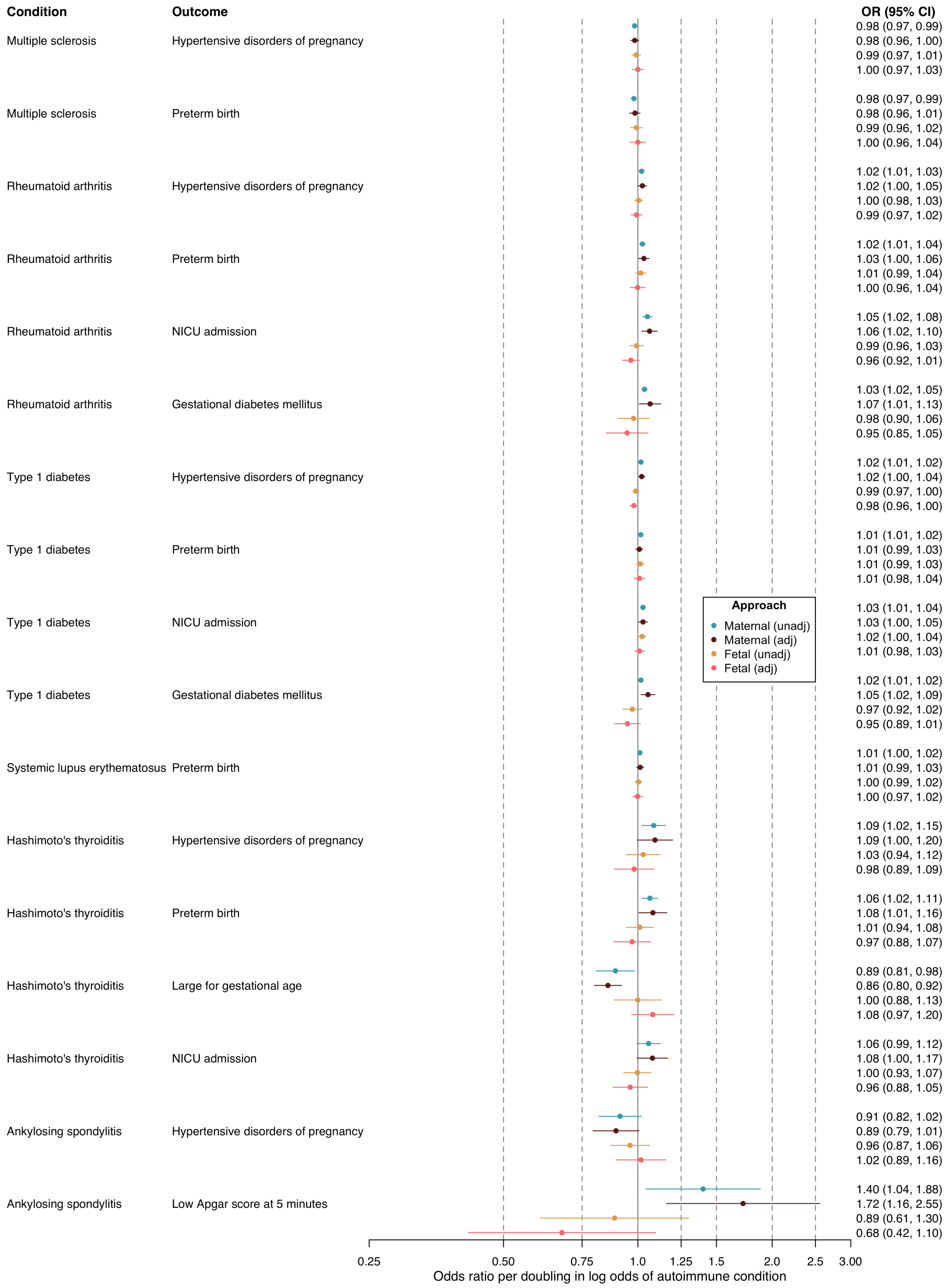

*Figure 5. Weighted linear model Mendelian randomization effect estimates exploring fetal genetic effects on the effects of increased log odds of autoimmune conditions on pregnancy outcomes, for results selected for follow up. Estimates reflect odds ratio of pregnancy outcome per doubling in log odds of autoimmune condition. Models shown are 1) maternal effects (unadjusted; same as main analysis inverse-variance weighted (IVW) estimates), 2) maternal effects adjusted for fetal genotype, 3) fetal effects (unadjusted), and 4) fetal effects adjusted for maternal genotype. Estimates reflect odds ratio of pregnancy outcome per doubling in log odds of autoimmune condition for mother (models 1 & 2) or fetus (models 3 & 4).*

*NICU = neonatal intensive care unit. The effect of type 1 diabetes liability on stillbirth is not included here since fetal genotypes are not available for this outcome.*

#### Leave one study out

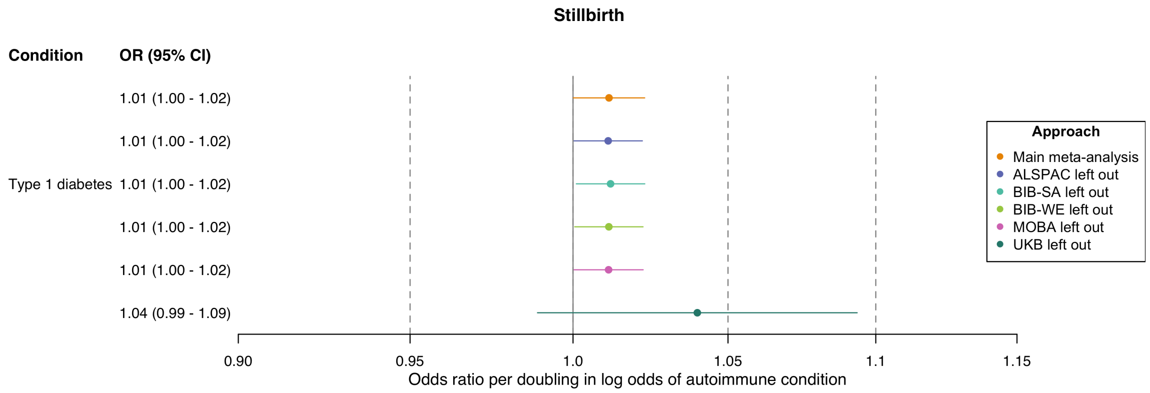

*Figure 6. Leave-one-study-out and main meta-analysis Mendelian randomization effect estimates for effects of increased log odds of autoimmune conditions on stillbirth, for results selected for follow up. Estimates reflect odds ratio of pregnancy outcome per doubling in log odds of autoimmune condition.*

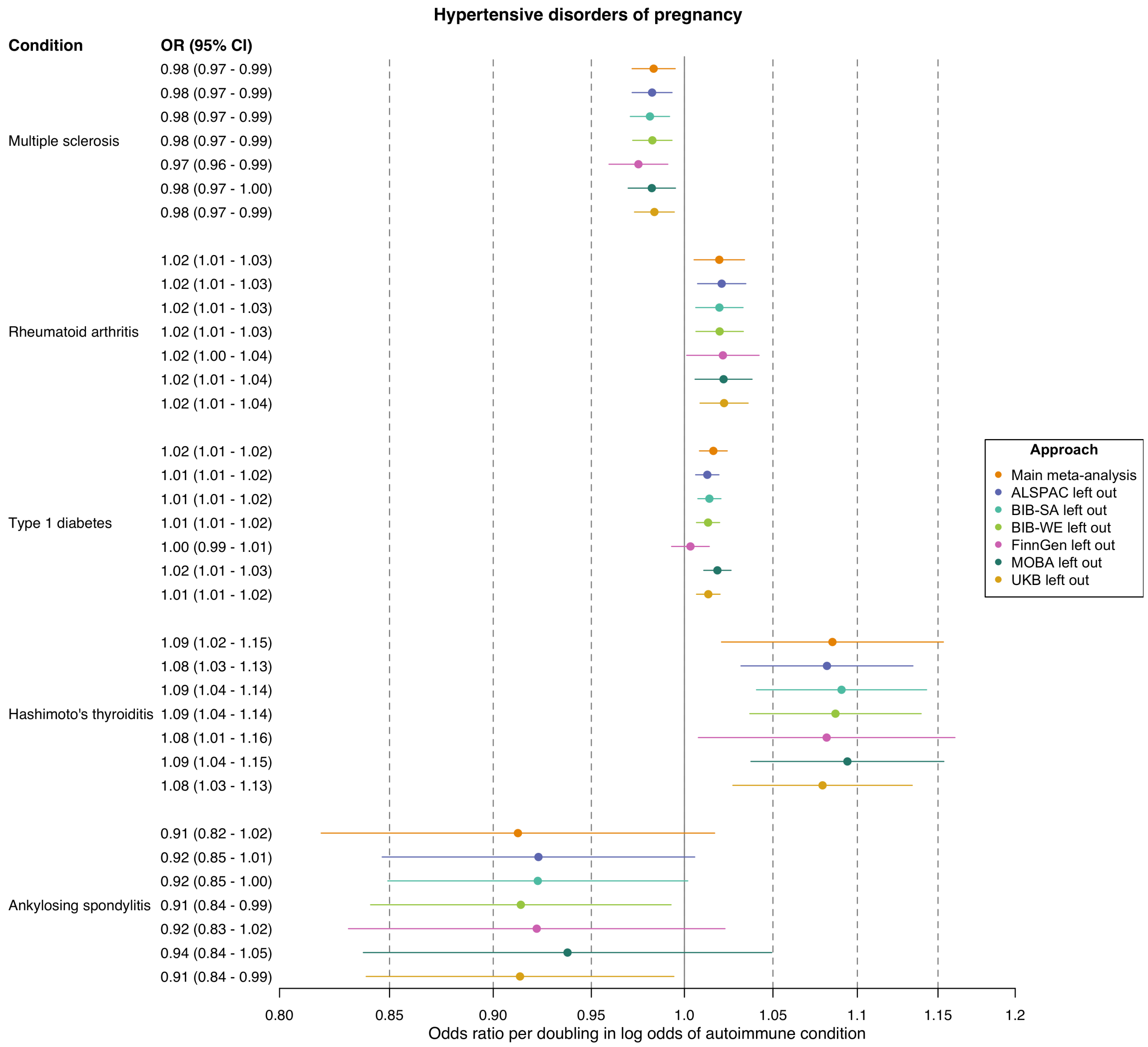

*Figure 7. Leave-one-study-out and main meta-analysis Mendelian randomization effect estimates for effects of increased log odds of autoimmune conditions on hypertensive disorders of pregnancy, for results selected for follow up. Estimates reflect odds ratio of pregnancy outcome per doubling in log odds of autoimmune condition.*

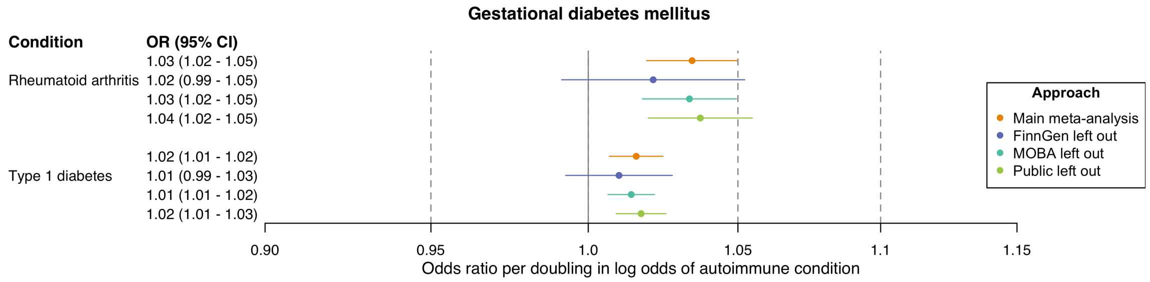

*Figure 8. Leave-one-study-out and main meta-analysis Mendelian randomization effect estimates for effects of increased log odds of autoimmune conditions on gestational diabetes mellitus, for results selected for follow up. Estimates reflect odds ratio of pregnancy outcome per doubling in log odds of autoimmune condition.*

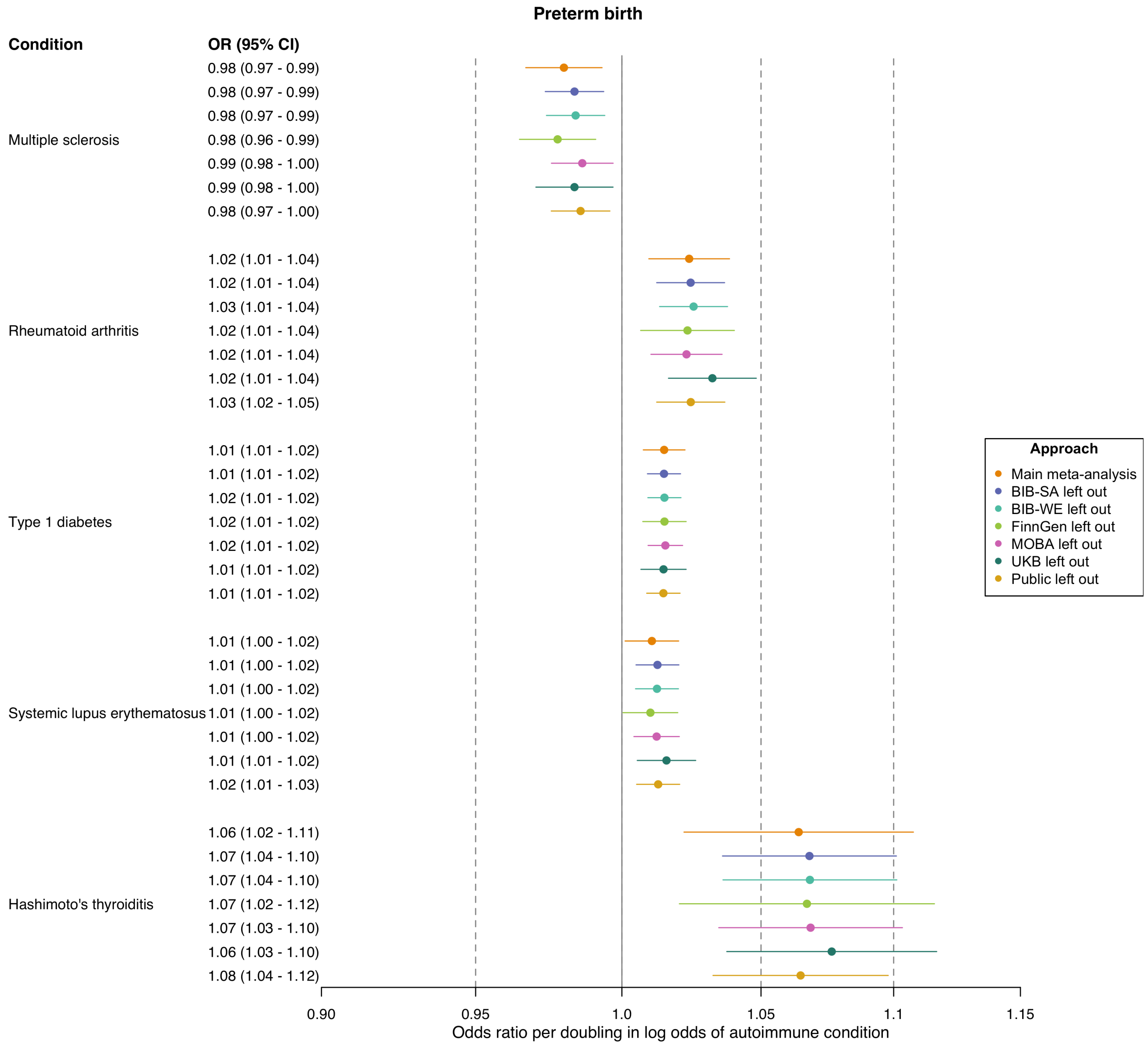

*Figure 9. Leave-one-study-out and main meta-analysis Mendelian randomization effect estimates for effects of increased log odds of autoimmune conditions on preterm birth, for results selected for follow up. Estimates reflect odds ratio of pregnancy outcome per doubling in log odds of autoimmune condition.*

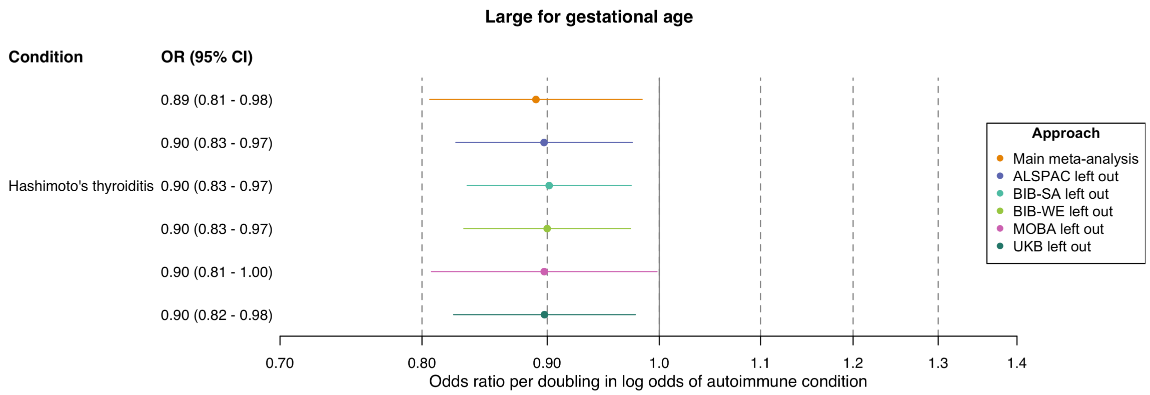

*Figure 10. Leave-one-study-out and main meta-analysis Mendelian randomization effect estimates for effects of increased log odds of autoimmune conditions on large for gestational age, for results selected for follow up. Estimates reflect odds ratio of pregnancy outcome per doubling in log odds of autoimmune condition.*

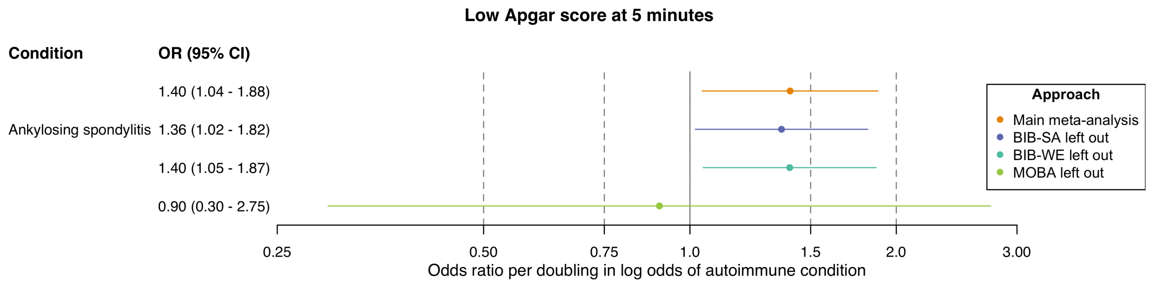

*Figure 11. Leave-one-study-out and main meta-analysis Mendelian randomization effect estimates for effects of increased log odds of autoimmune conditions on low Apgar score at 5 minutes, for results selected for follow up. Estimates reflect odds ratio of pregnancy outcome per doubling in log odds of autoimmune condition.*

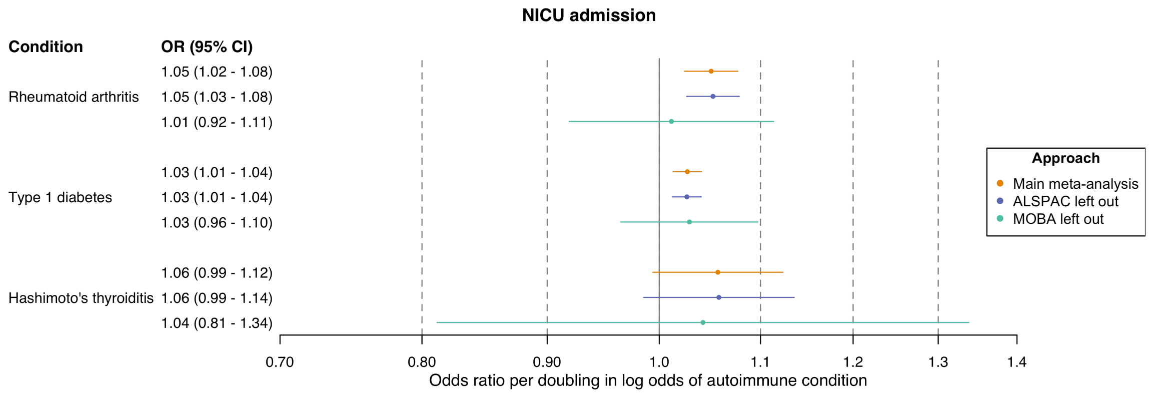

*Figure 12. Leave-one-study-out and main meta-analysis Mendelian randomization effect estimates for effects of increased log odds of autoimmune conditions on NICU admission, for results selected for follow up. Estimates reflect odds ratio of pregnancy outcome per doubling in log odds of autoimmune condition. NICU = neonatal intensive care unit.*

### Leave one SNP out estimates

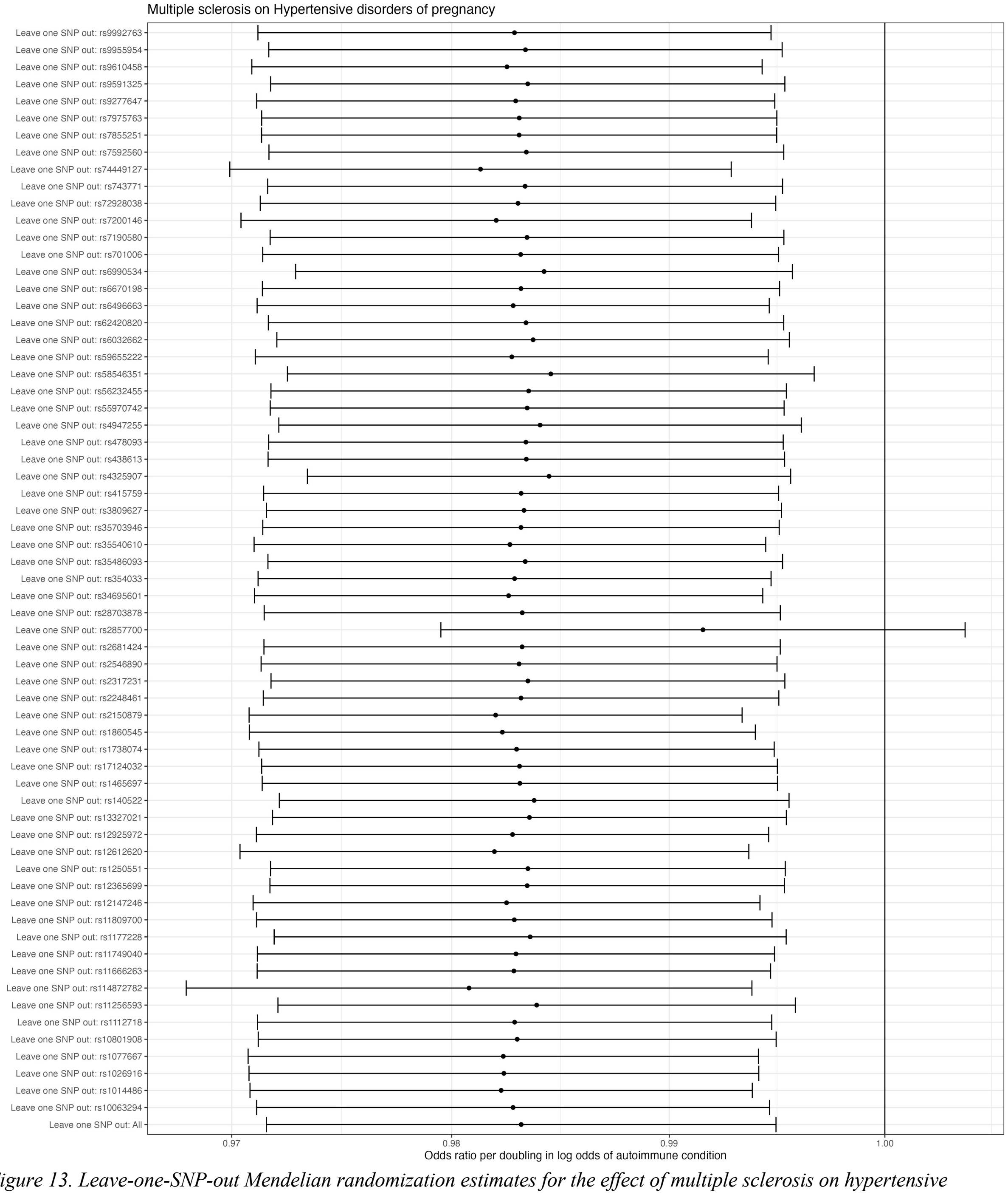

*Figure 13. Leave-one-SNP-out Mendelian randomization estimates for the effect of multiple sclerosis on hypertensive disorders of pregnancy. Estimates reflect odds ratio of pregnancy outcome per doubling in log odds of autoimmune condition.*

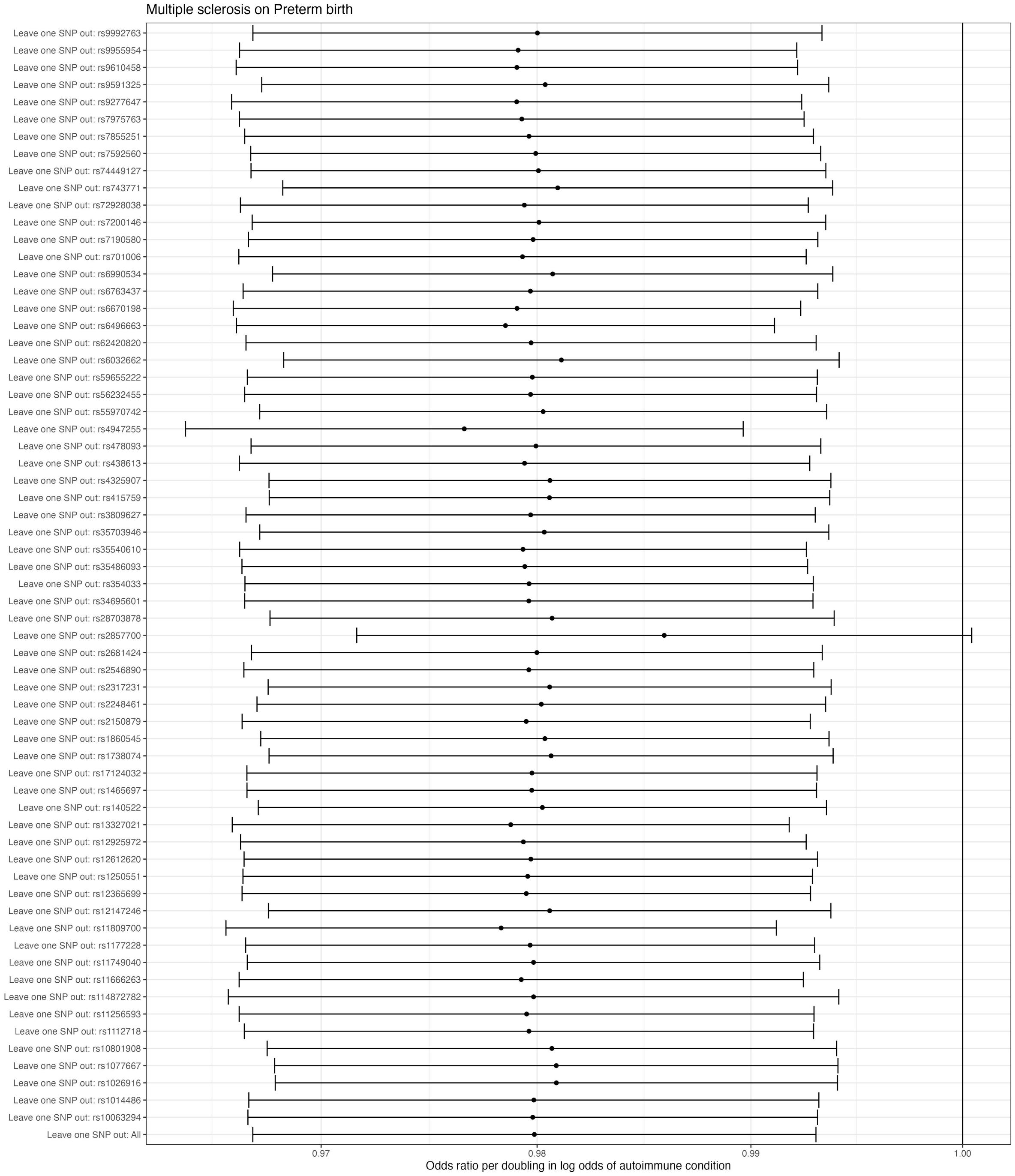

*Figure 14. Leave-one-SNP-out Mendelian randomization estimates for the effect of multiple sclerosis on preterm birth. Estimates reflect odds ratio of pregnancy outcome per doubling in log odds of autoimmune condition.*

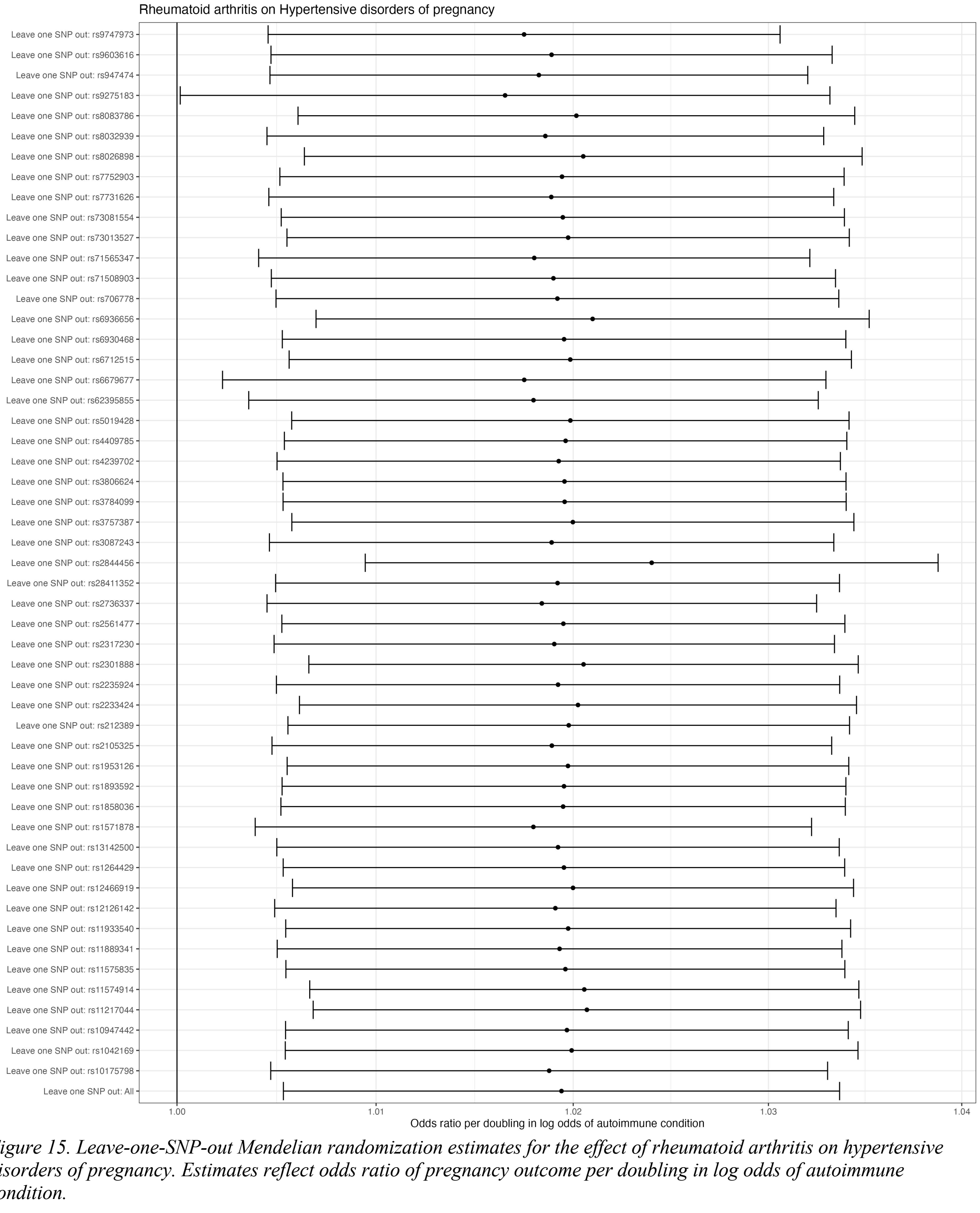

*Figure 15. Leave-one-SNP-out Mendelian randomization estimates for the effect of rheumatoid arthritis on hypertensive disorders of pregnancy. Estimates reflect odds ratio of pregnancy outcome per doubling in log odds of autoimmune condition.*

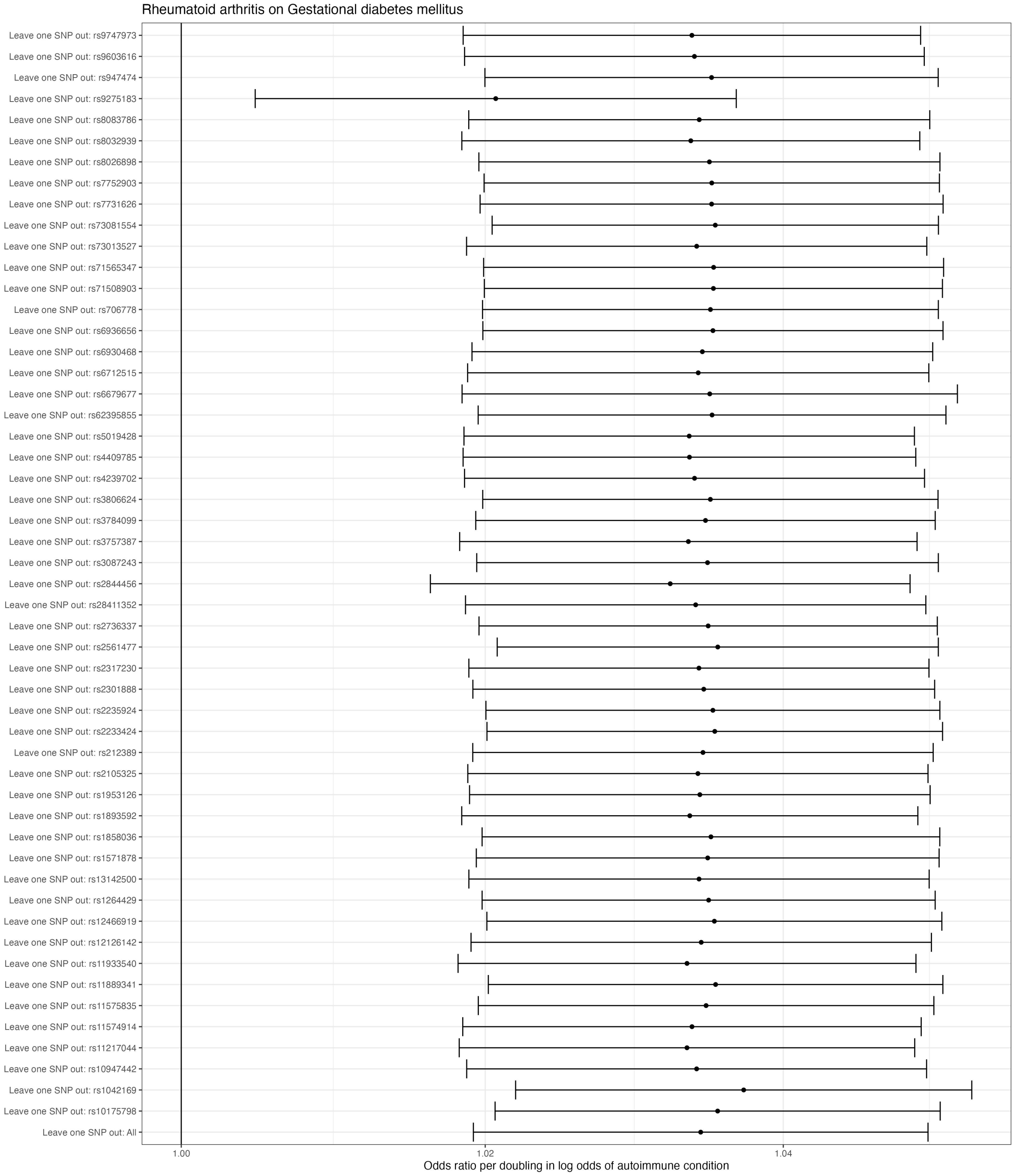

*Figure 16. Leave-one-SNP-out Mendelian randomization estimates for the effect of rheumatoid arthritis on gestational diabetes mellitus. Estimates reflect odds ratio of pregnancy outcome per doubling in log odds of autoimmune condition.*

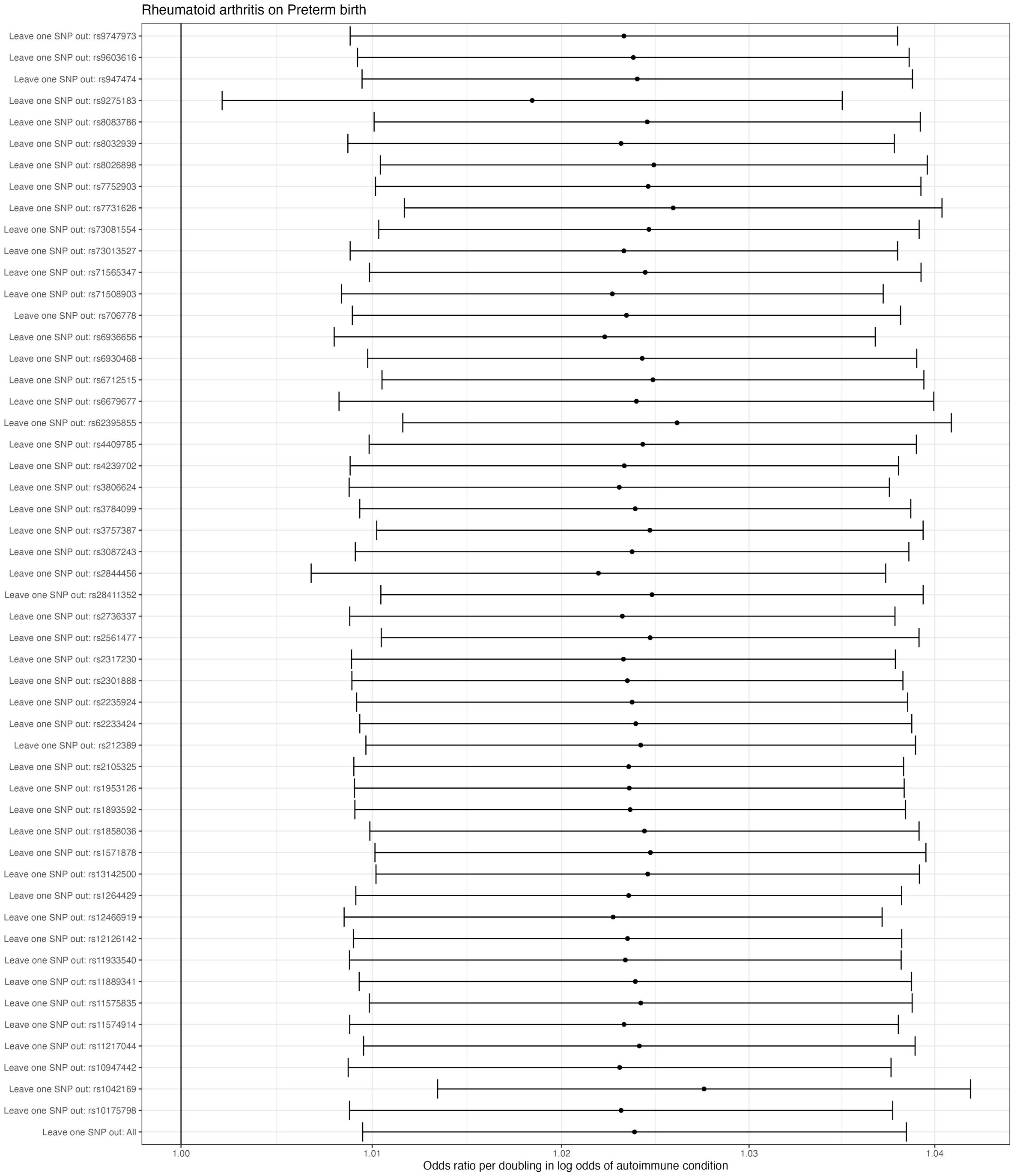

*Figure 17. Leave-one-SNP-out Mendelian randomization estimates for the effect of rheumatoid arthritis on preterm birth. Estimates reflect odds ratio of pregnancy outcome per doubling in log odds of autoimmune condition.*

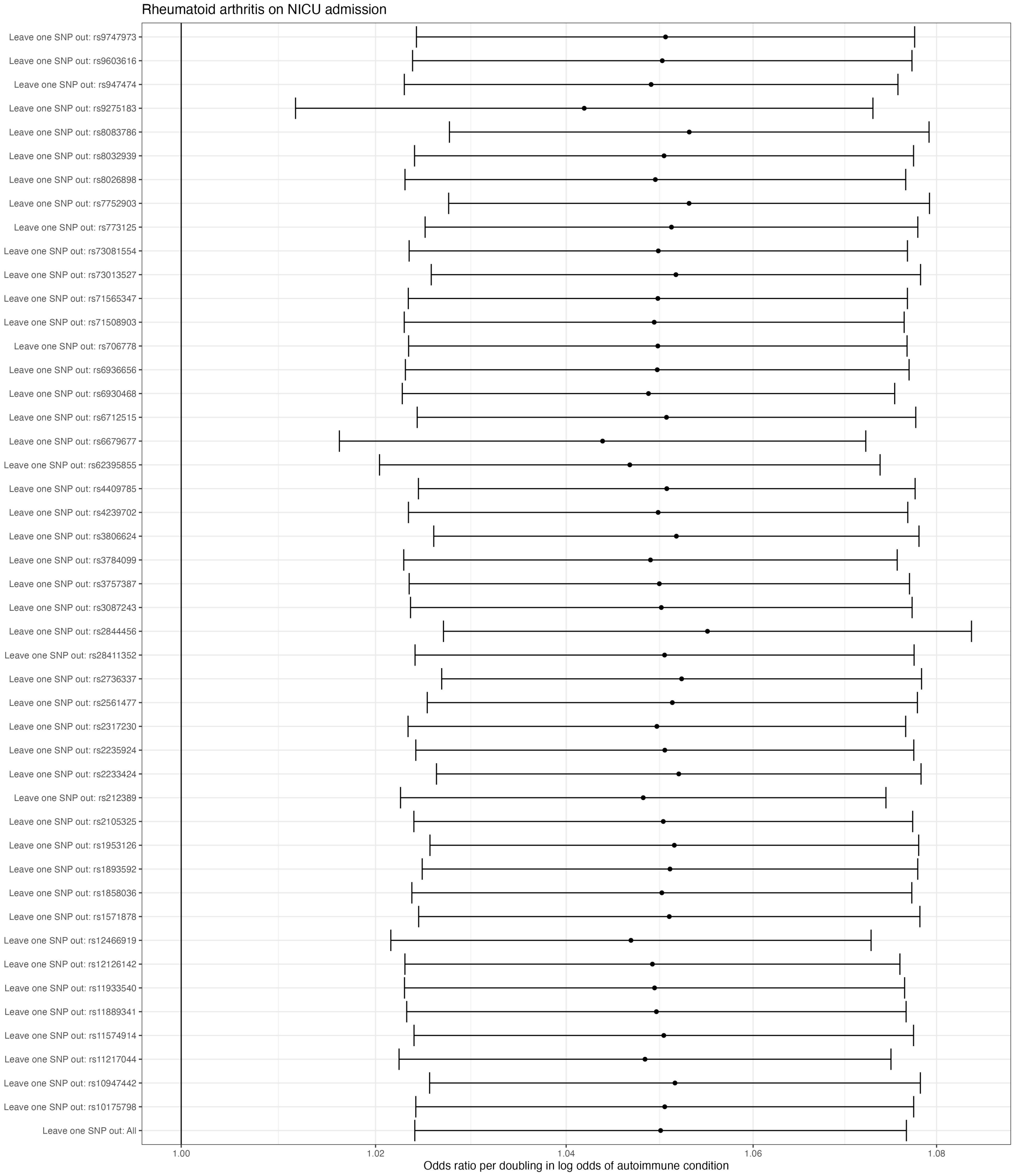

*Figure 18. Leave-one-SNP-out Mendelian randomization estimates for the effect of rheumatoid arthritis on NICU admission. Estimates reflect odds ratio of pregnancy outcome per doubling in log odds of autoimmune condition.*

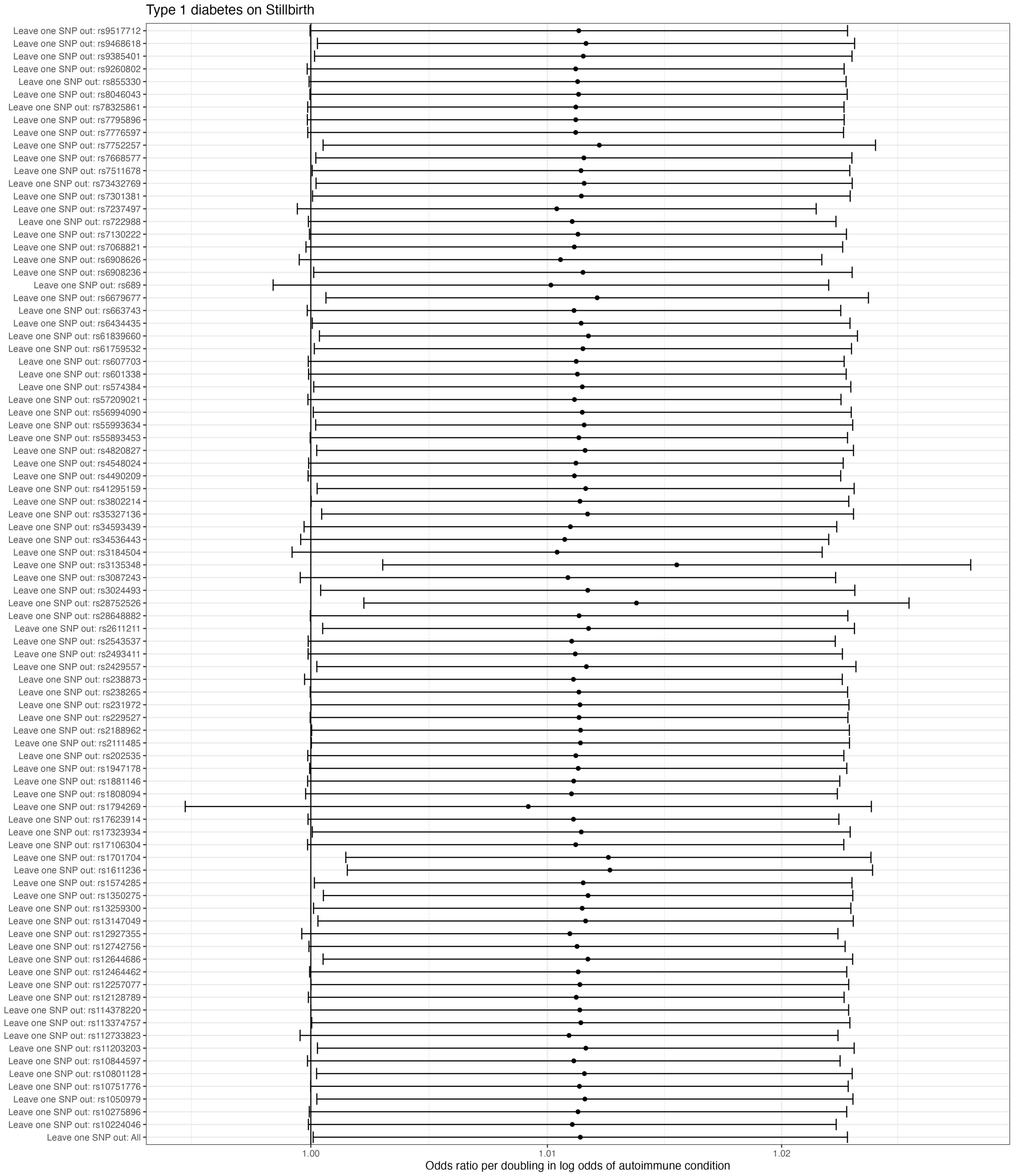

*Figure 19. Leave-one-SNP-out Mendelian randomization estimates for the effect of type 1 diabetes on stillbirth. Estimates reflect odds ratio of pregnancy outcome per doubling in log odds of autoimmune condition.*

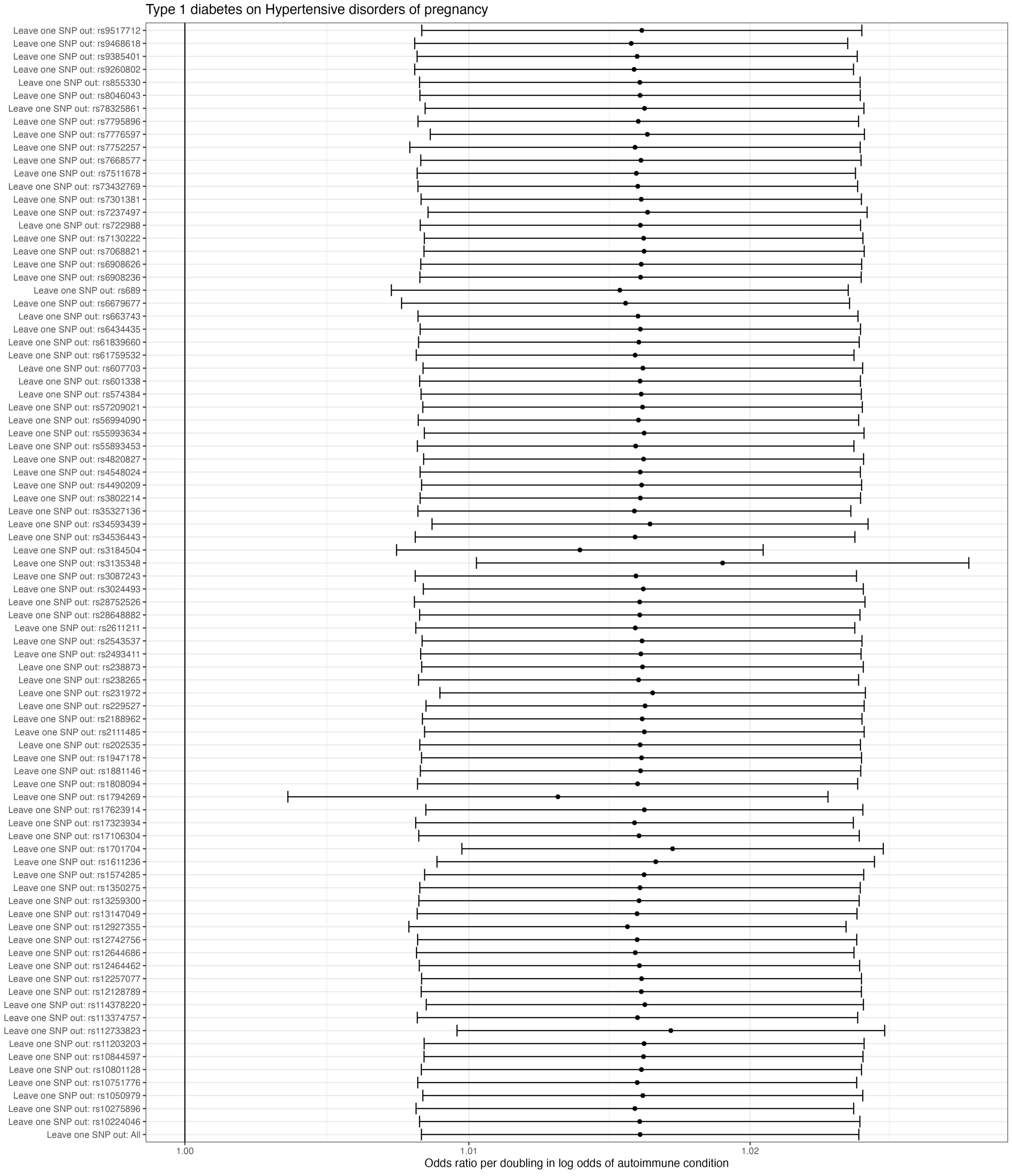

*Figure 20. Leave-one-SNP-out Mendelian randomization estimates for the effect of type 1 diabetes on hypertensive disorders of pregnancy. Estimates reflect odds ratio of pregnancy outcome per doubling in log odds of autoimmune condition.*

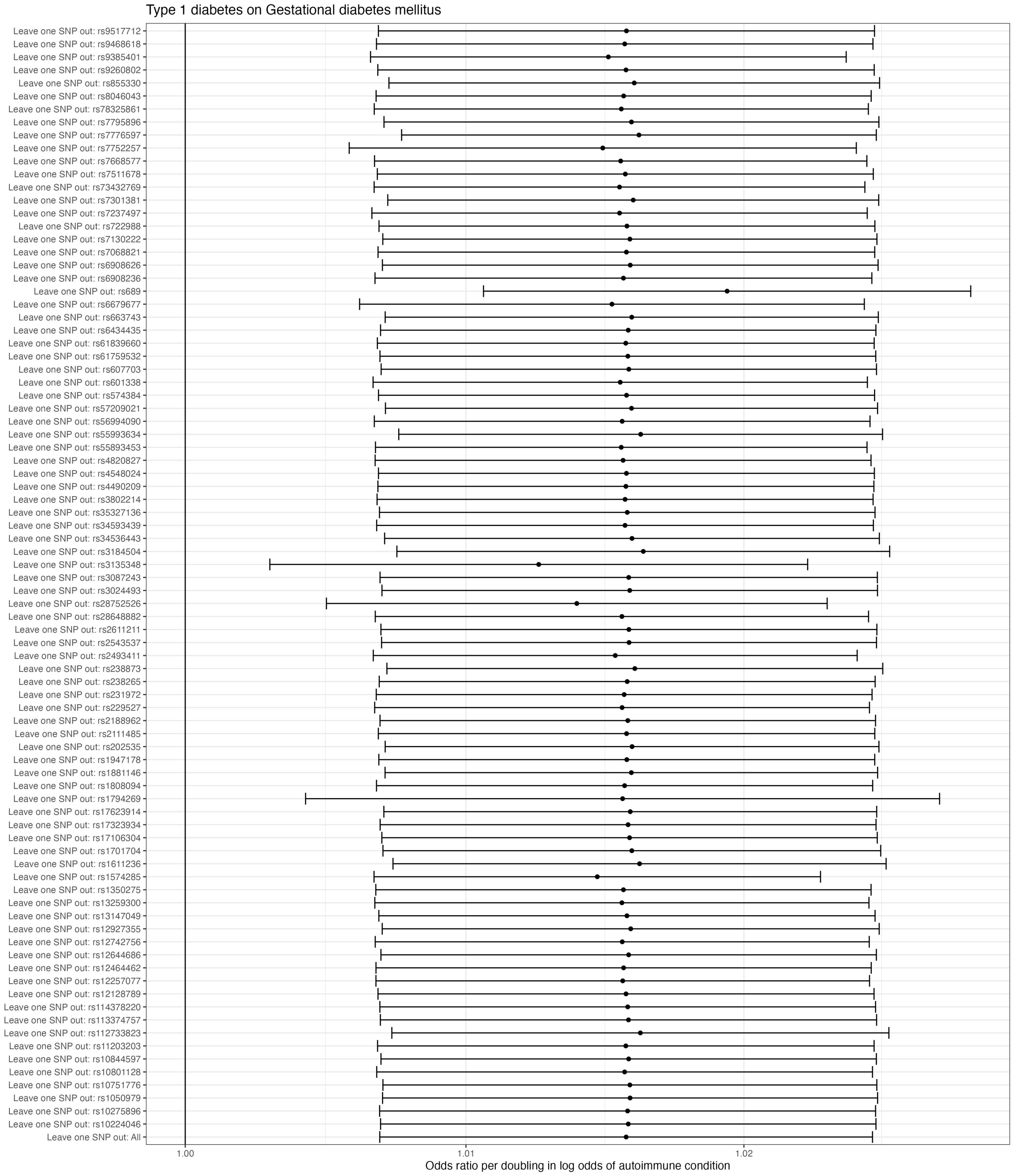

*Figure 21. Leave-one-SNP-out Mendelian randomization estimates for the effect of type 1 diabetes on gestational diabetes mellitus. Estimates reflect odds ratio of pregnancy outcome per doubling in log odds of autoimmune condition.*

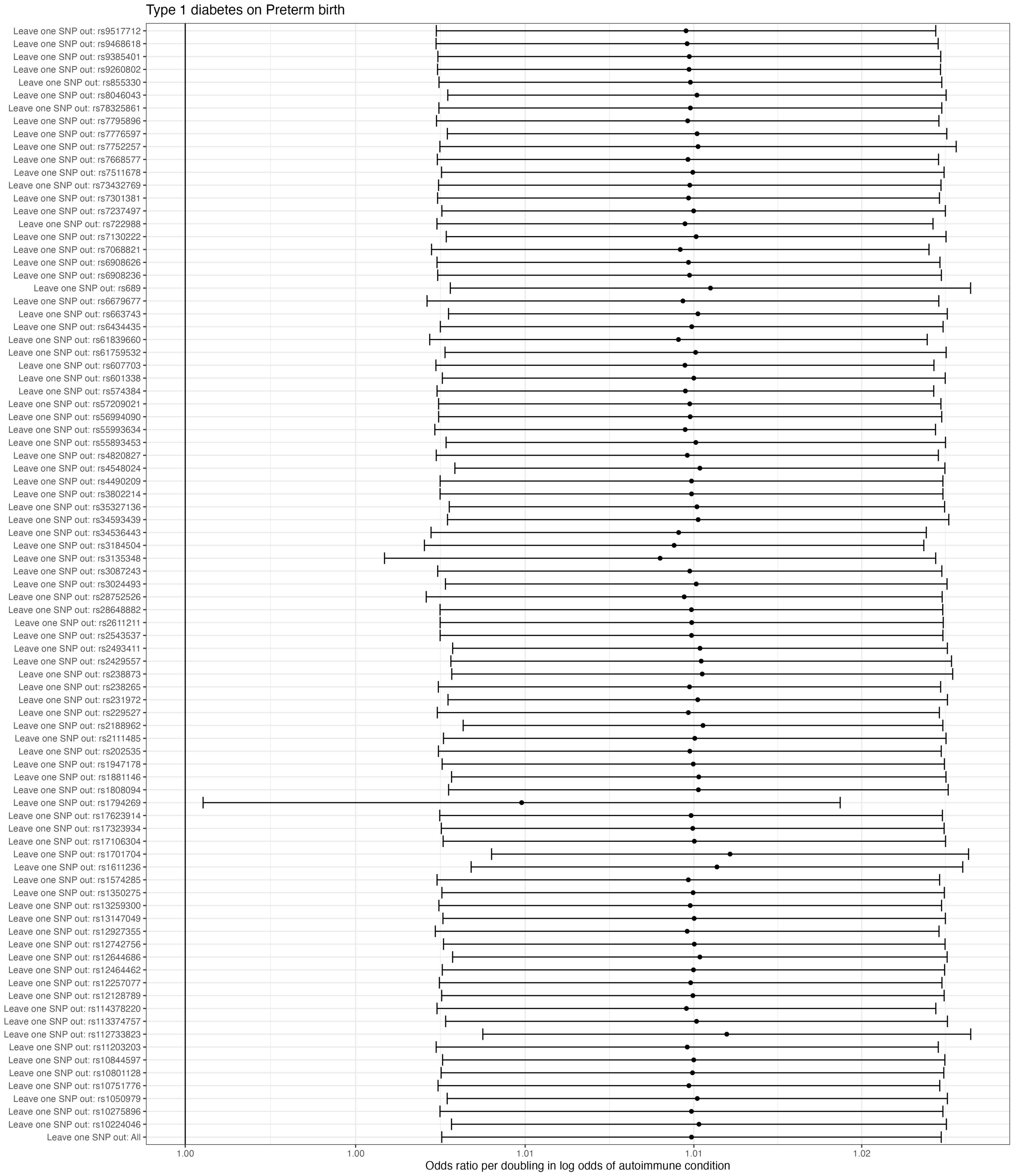

*Figure 22. Leave-one-SNP-out Mendelian randomization estimates for the effect of type 1 diabetes on preterm birth. Estimates reflect odds ratio of pregnancy outcome per doubling in log odds of autoimmune condition.*

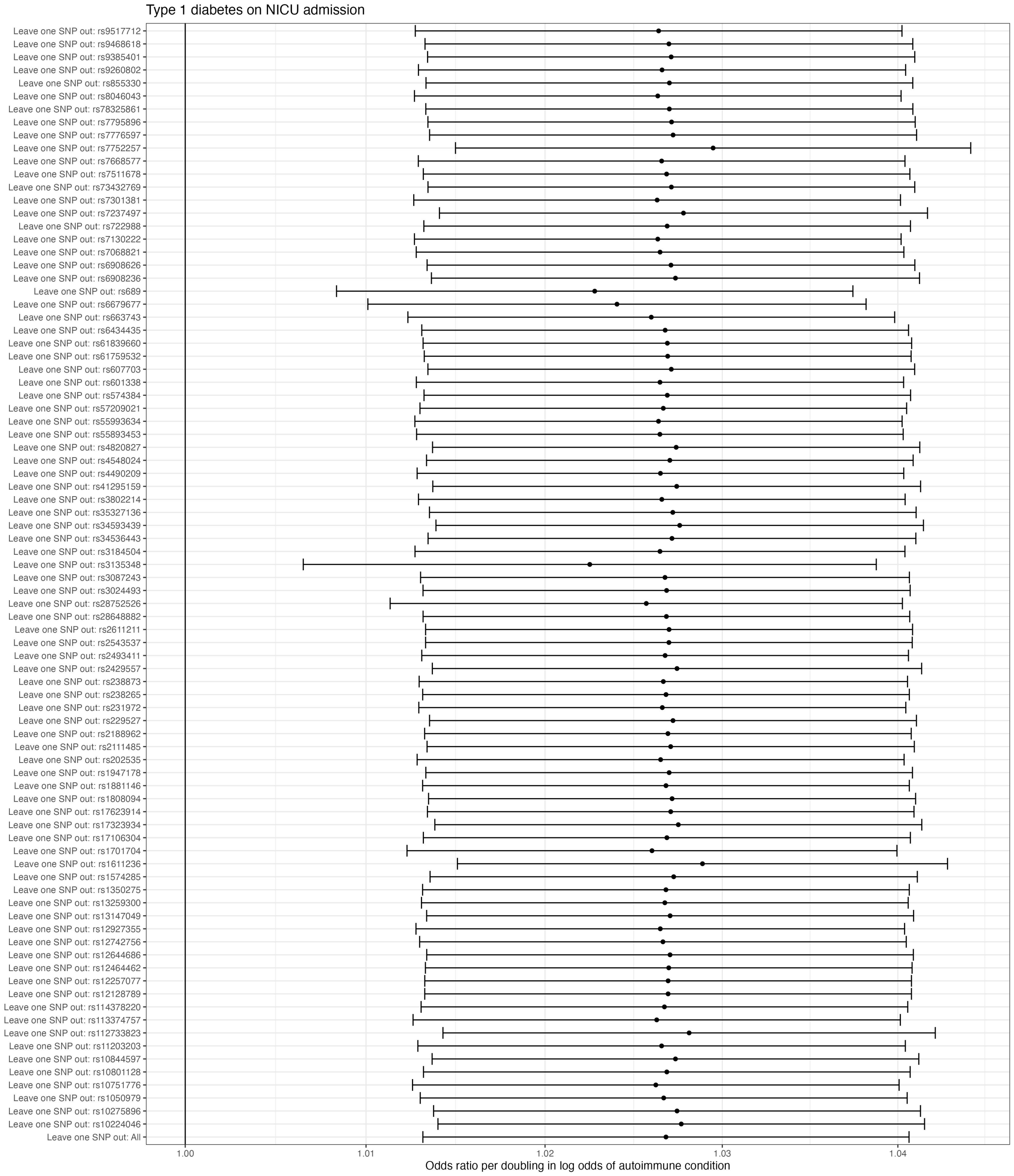

*Figure 23. Leave-one-SNP-out Mendelian randomization estimates for the effect of type 1 diabetes on NICU admission. Estimates reflect odds ratio of pregnancy outcome per doubling in log odds of autoimmune condition.*

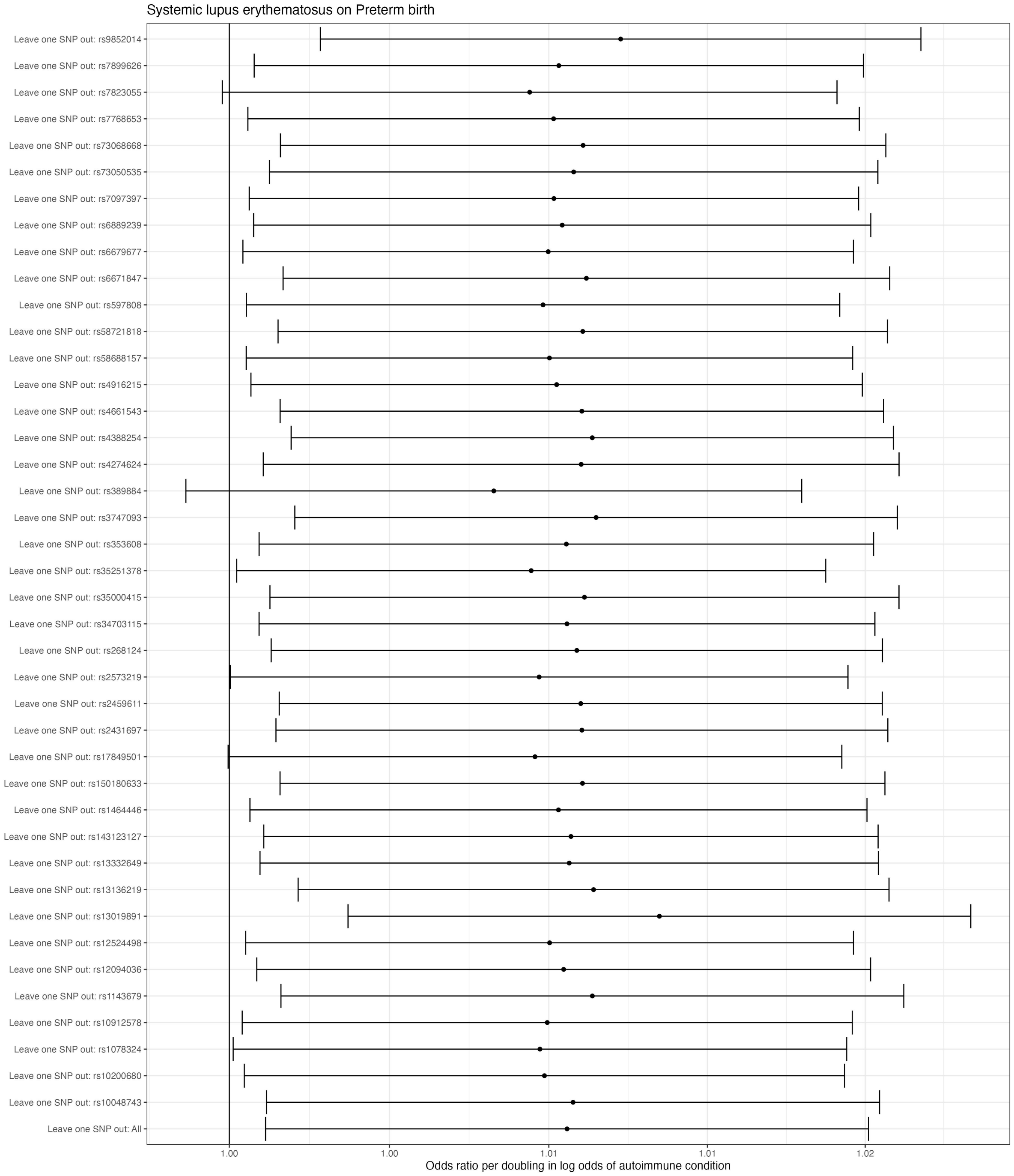

*Figure 24. Leave-one-SNP-out Mendelian randomization estimates for the effect of systemic lupus erythematosus on preterm birth. Estimates reflect odds ratio of pregnancy outcome per doubling in log odds of autoimmune condition.*

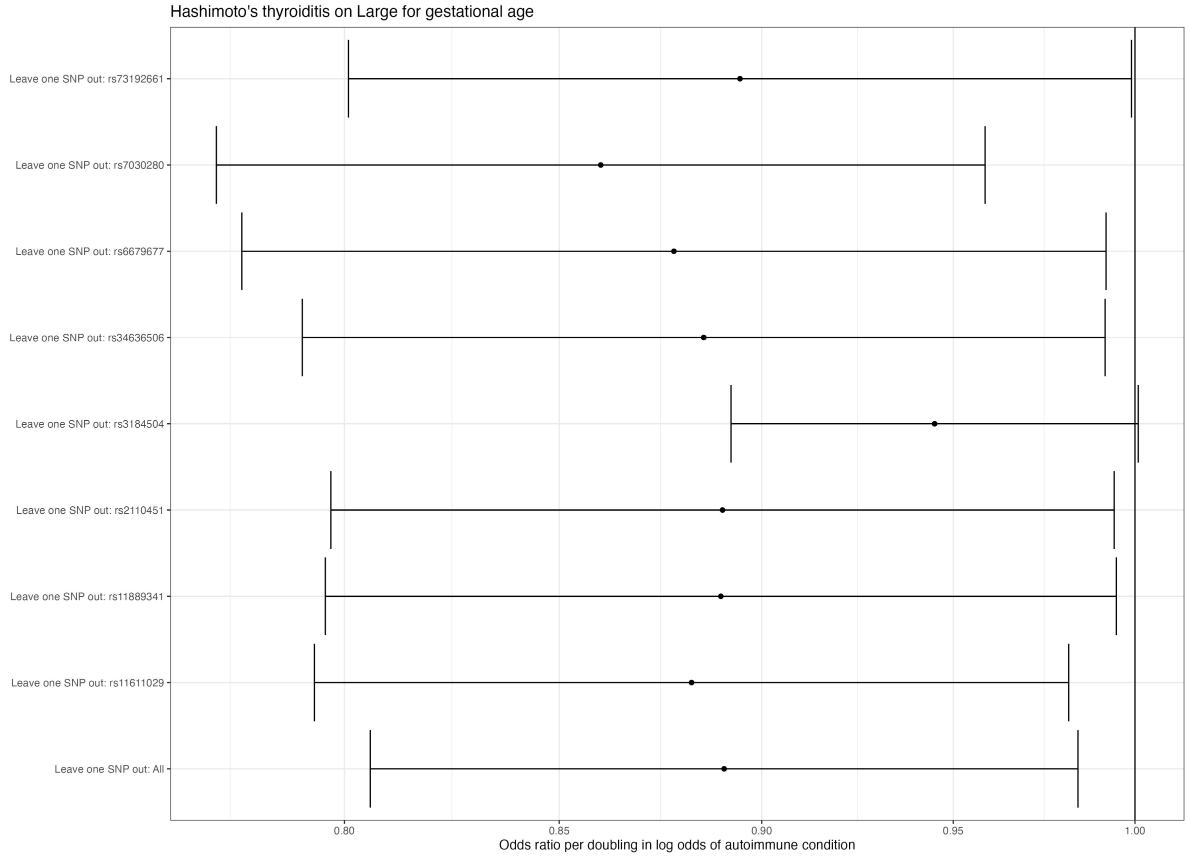

*Figure 25. Leave-one-SNP-out Mendelian randomization estimates for the effect of Hashimoto's thyroiditis on large-for-gestational-age. Estimates reflect odds ratio of pregnancy outcome per doubling in log odds of autoimmune condition.*

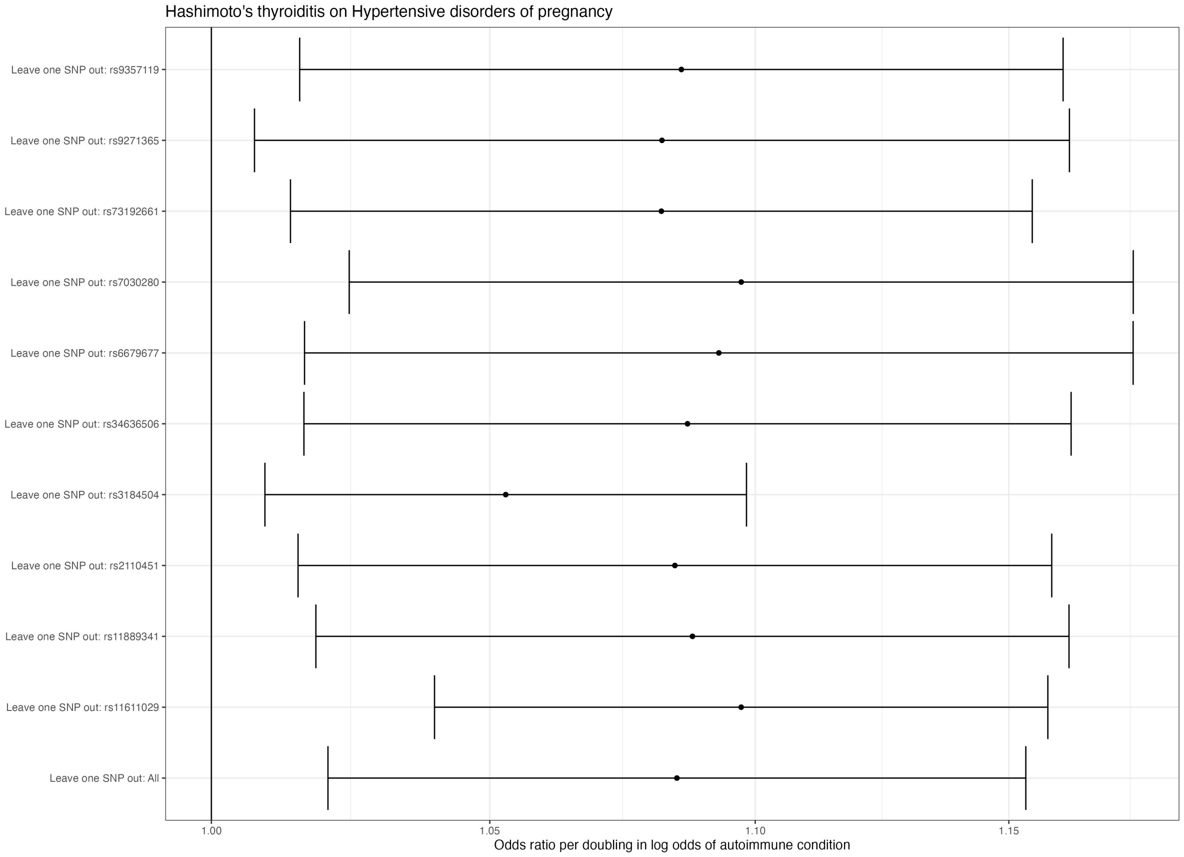

*Figure 26. Leave-one-SNP-out Mendelian randomization estimates for the effect of Hashimoto's thyroiditis on hypertensive disorders of pregnancy. Estimates reflect odds ratio of pregnancy outcome per doubling in log odds of autoimmune condition.*

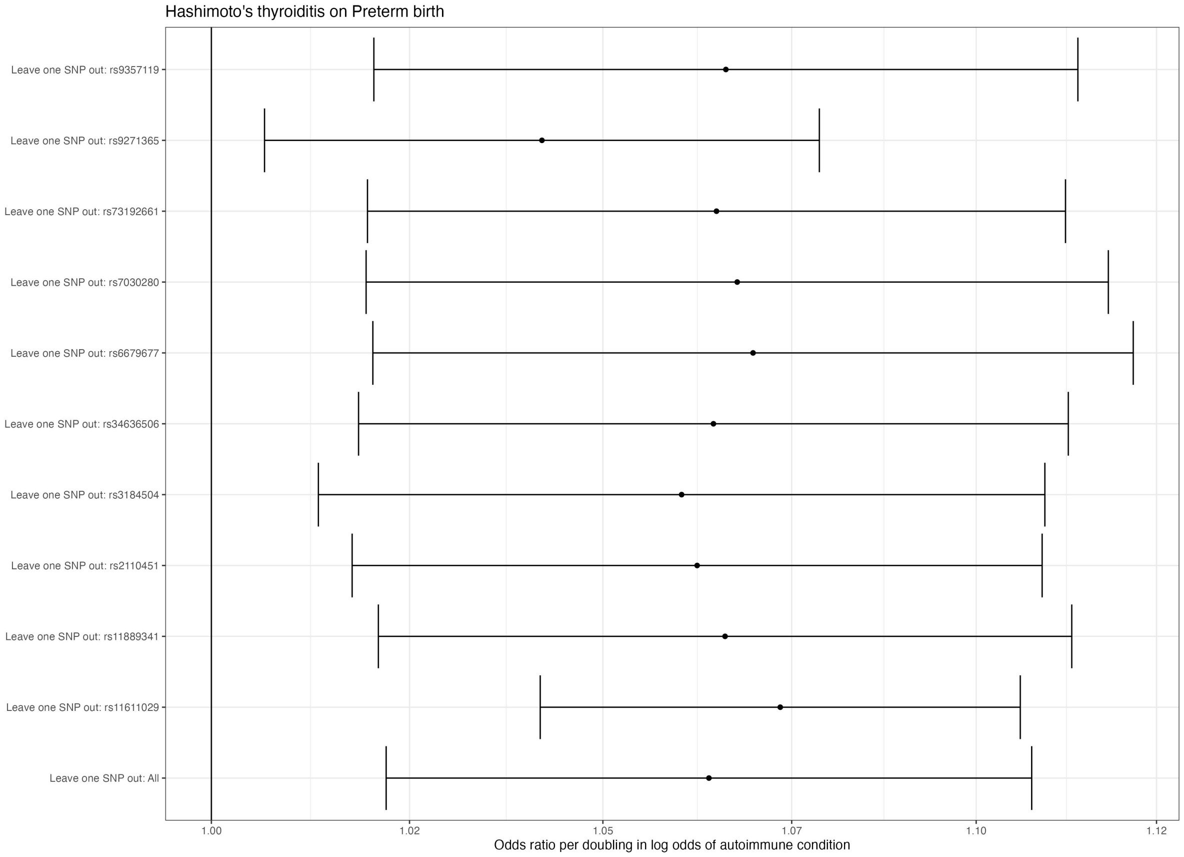

*Figure 27. Leave-one-SNP-out Mendelian randomization estimates for the effect of Hashimoto's thyroiditis on preterm birth. Estimates reflect odds ratio of pregnancy outcome per doubling in log odds of autoimmune condition.*

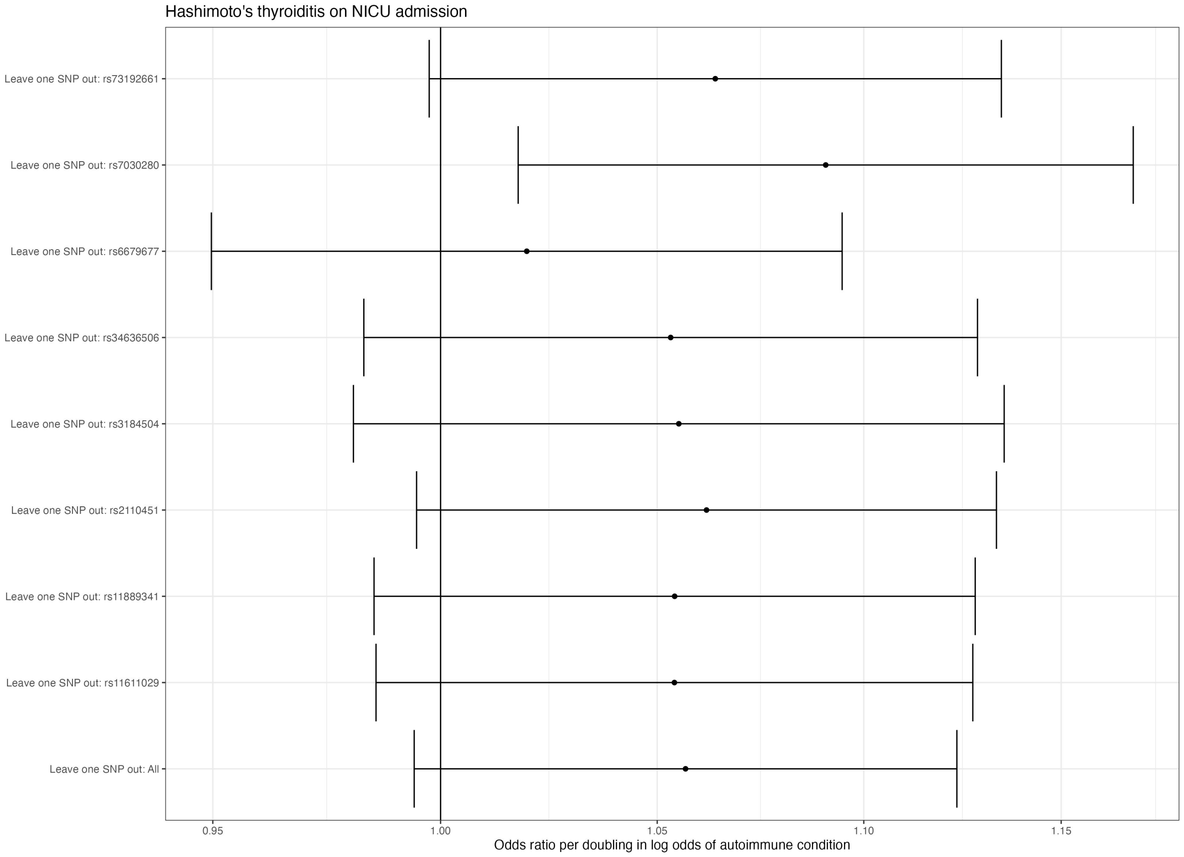

*Figure 28. Leave-one-SNP-out Mendelian randomization estimates for the effect of Hashimoto's thyroiditis on NICU admission. Estimates reflect odds ratio of pregnancy outcome per doubling in log odds of autoimmune condition.*

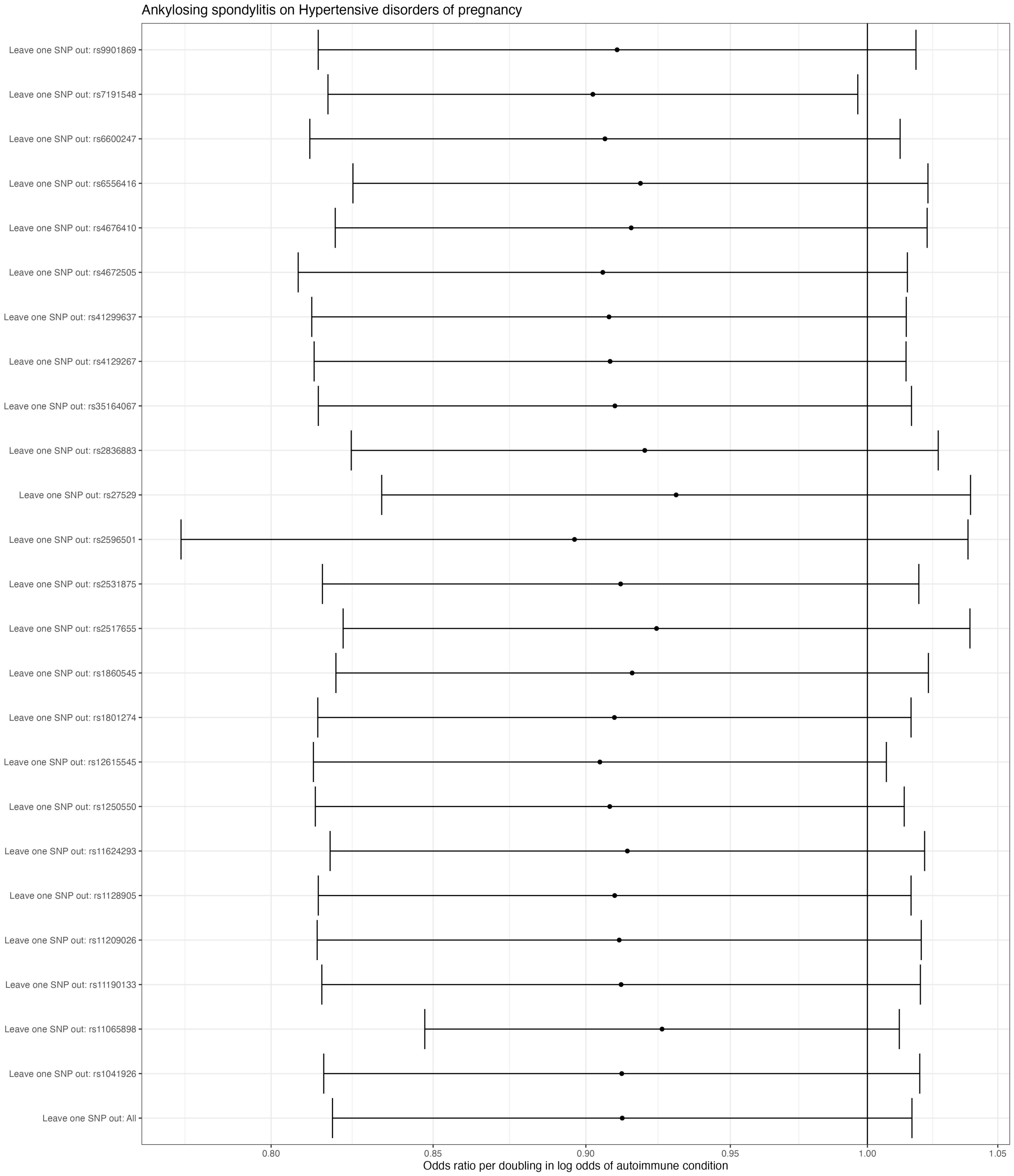

*Figure 29. Leave-one-SNP-out Mendelian randomization estimates for the effect of ankylosing spondylitis on hypertensive disorders of pregnancy. Estimates reflect odds ratio of pregnancy outcome per doubling in log odds of autoimmune condition.*

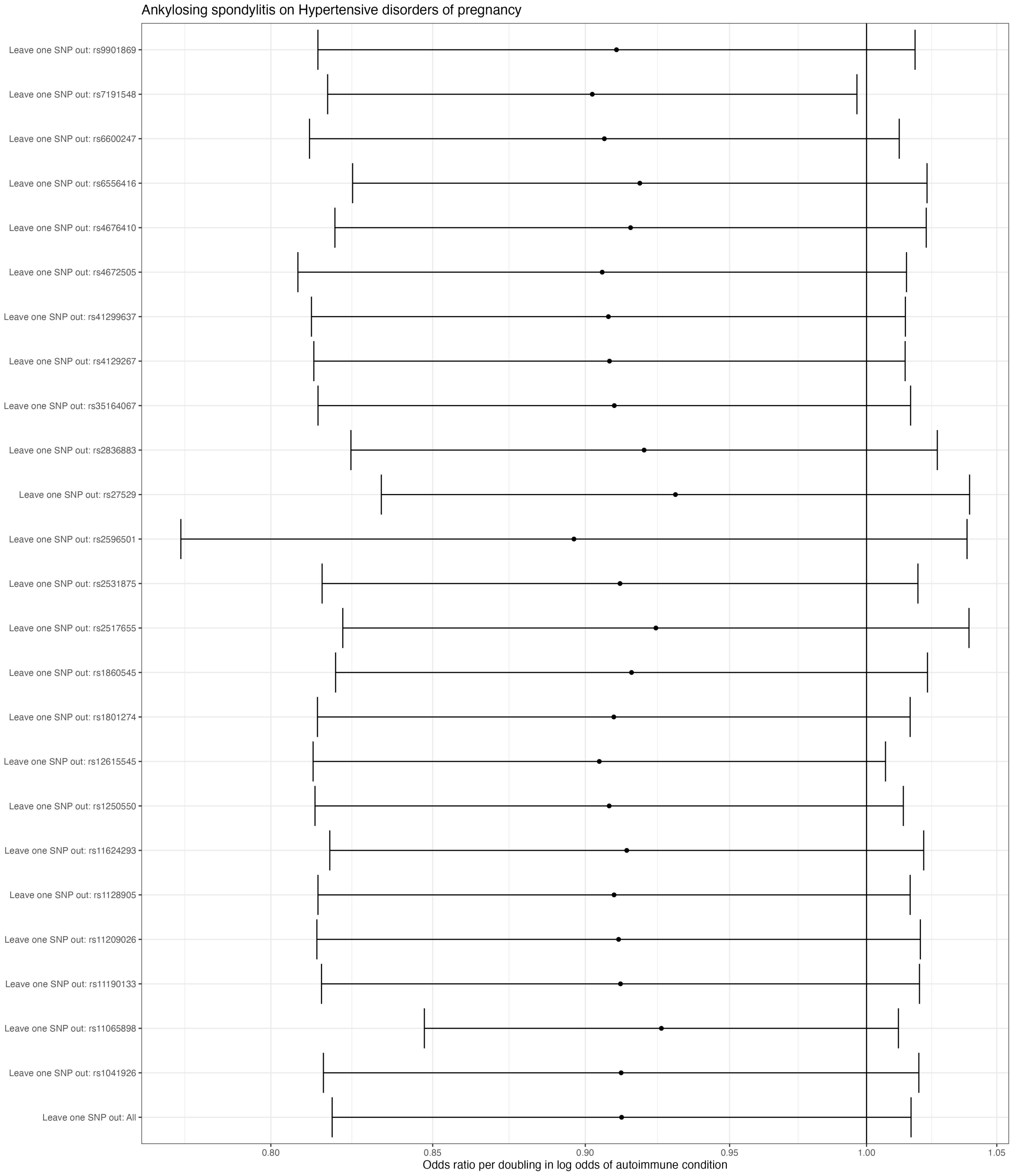

*Figure 30. Leave-one-SNP-out Mendelian randomization estimates for the effect of ankylosing spondylitis on low Apgar score at 5 minutes. Estimates reflect odds ratio of pregnancy outcome per doubling in log odds of autoimmune condition.*

#### Phenome wide scan for influential SNPs

**rs2857700**

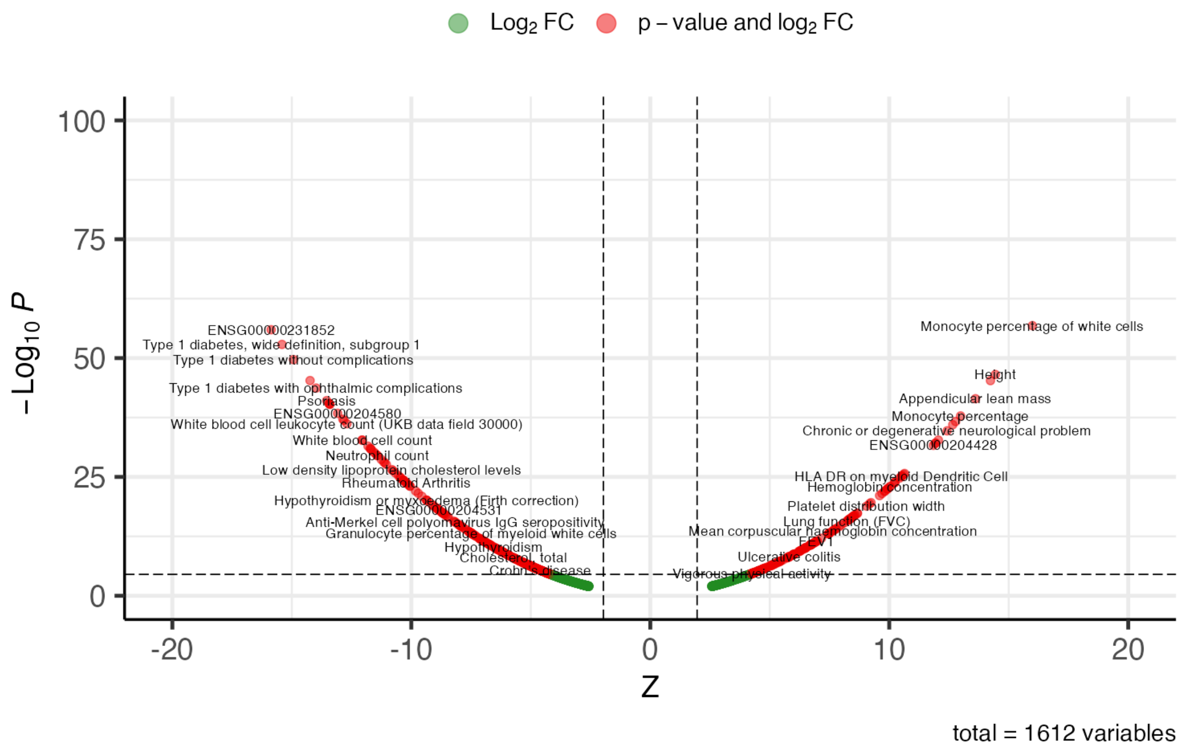

*Figure 31. Phenome scan results for rs2857700, instrumenting multiple sclerosis.*

**rs9275183**

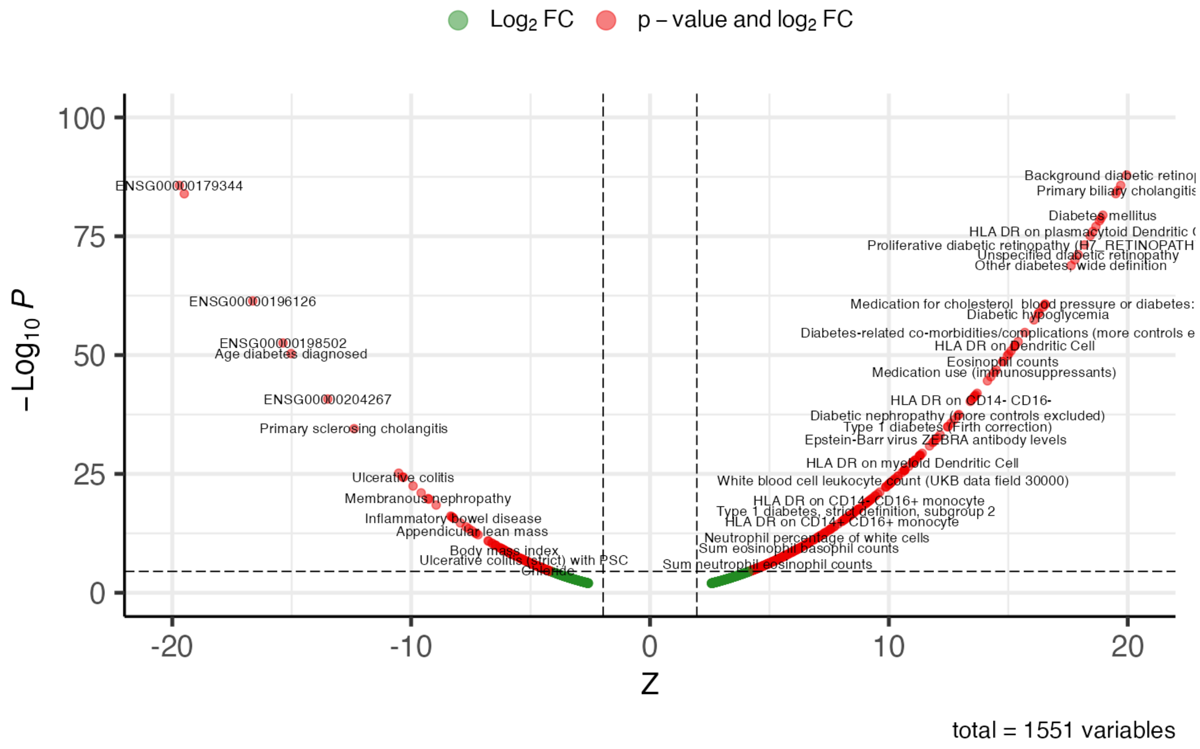

*Figure 32. PheWAS results for rs9275183, instrumenting rheumatoid arthritis.*

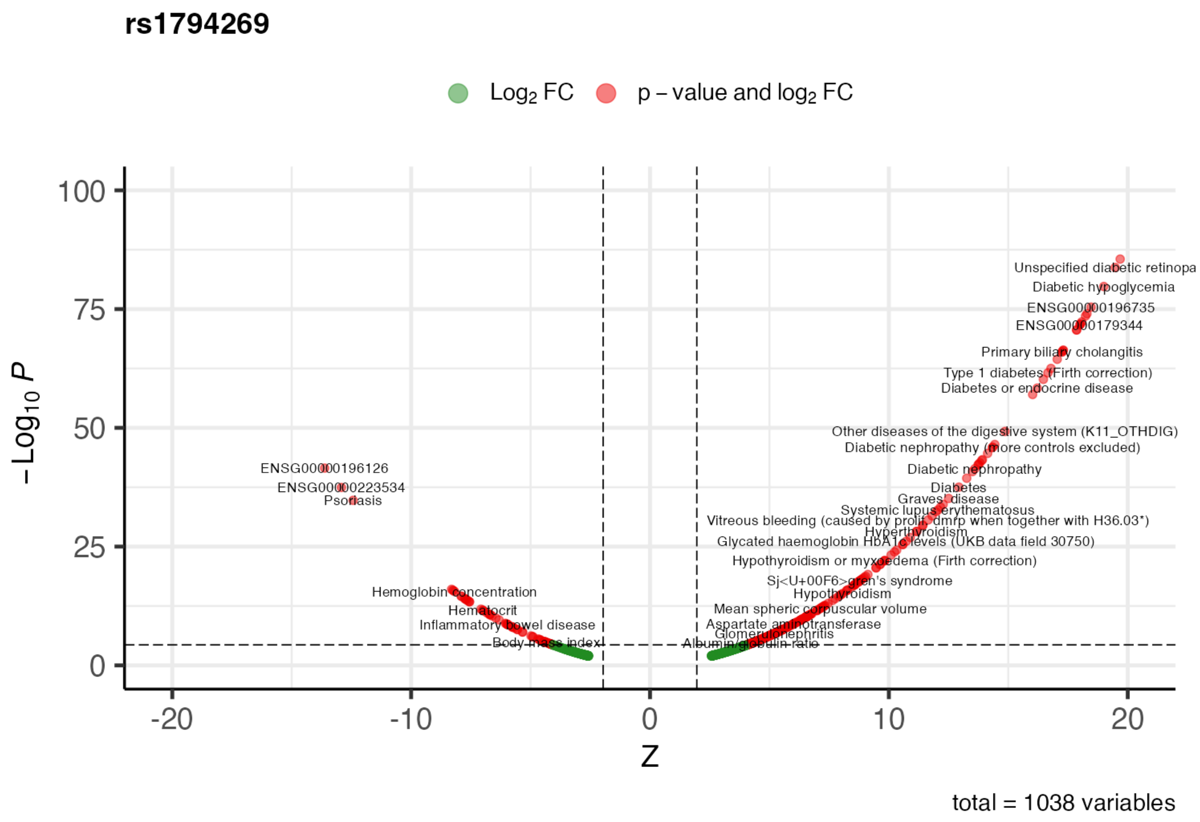

*Figure 33. PheWAS results for rs1794269, instrumenting type 1 diabetes.*

#### Instrument strength

We estimated instrument strength using the equation:

$$F = \left(\frac{beta}{standard\ error}\right)^2$$

This form was used instead of versions requiring effect allele frequency information since this was not available for all genetic variants in the selected exposure GWAS.

#### Fetal genetic effects

To estimate effects of maternal genetic instruments on outcomes while accounting for fetal genetic effects, we used a weighted linear model (WLM). The WLM enables estimation of the mutually adjusted effects of maternal and fetal genotype on the outcomes, which are calculated using the summary-level association data from unadjusted GWAS (in which the outcomes were regressed on maternal or fetal genotype separately)[1–4]. Unlike a traditional approach, in which the outcome is regressed on maternal and fetal genotype jointly in a sample with both maternal and fetal genotype data available, the unadjusted GWAS used for the WLM require only maternal or fetal genotype data and can be fitted on separate (potentially overlapping) samples. This enables more efficient use of the available data.

To generate the WLM adjusted genetic estimates within the MR-PREG collaboration, we first generated unadjusted GWAS by regressing the outcomes separately on maternal and fetal genotype (and covariates including genetic principal components[5]. The MR-PREG unadjusted maternal GWAS used data from five cohort studies and two publicly available GWAS consortium datasets. The MR-PREG unadjusted fetal GWAS used data from three cohort studies (ALSPAC, BiB and MoBa). We then implemented the WLM using the DONUTS R package[3], to estimate i) maternal associations adjusted for offspring genotype and ii) fetal associations adjusted for maternal genotype. These models accounted for sample overlap between the maternal and fetal GWAS using an intercept term from bivariate LD score regression[6] of the maternal and fetal GWAS. The underlying equations are outlined in the MR-PREG collaboration profile supplementary material[5].

In our study no exposure instruments for systemic sclerosis (n=12 SNPs) were available in the fetal adjusted outcome GWAS, so we were unable to conduct the sensitivity analysis adjusting for fetal genotype for systemic sclerosis.

#### Cohort descriptions

*Avon Longitudinal Study of Parents and Children (ALSPAC)*

Pregnant women resident in Avon, UK with expected dates of delivery between 1st April 1991 and 31st December 1992 were invited to take part in the study[7,8]. 20,248 pregnancies have been identified as being eligible and the initial number of pregnancies enrolled was 14,541 (ALSPAC-G0), and gave birth to a total of 14,676 foetuses (ALSPAC-G1). These resulted in 14,062 live births and 13,988 children who were alive at 1 year of age. The total sample size for analyses using any data collected after the age of seven is therefore 15,447 pregnancies, resulting in 15,658 foetuses. Of these 14,901 children were alive at 1 year of age. A total of 14,203 unique mothers were initially enrolled in the study, and after additional recruitment a total of 14,833 unique women (ALSPAC-G0 mothers) were enrolled in ALSPAC as of September 2021. Here, we integrate ALSPAC data on these G0 pregnancies from: genetic data for mothers and children, four questionnaires collected during pregnancy, and linked obstetric records.

*Born in Bradford (BiB)*

BiB prospectively recruited pregnant women in Bradford with expected dates of delivery between March 2007 and December 2010[9]. Recruitment for most women was at their oral glucose tolerance test, offered to all women booked for delivery at Bradford Royal Infirmary at around 26-28 weeks gestation. A total of 12,453 women (13,776 pregnancies) were enrolled during pregnancy, who gave birth to 13,858 live children. In this study, we used BiB genetic data collected from both mothers and children, and pregnancy outcome information retrieved through questionnaires and routinely linked primary and secondary care records.

*Norwegian Mother, Father and Child Cohort Study (MoBa)*

MoBa prospectively recruited pregnant women across Norway between 1999 and 2008. The cohort has recruited approximately 95,200 mothers, 75,200 fathers and 114,500 children. Here, we used genetic data for mothers and children, alongside questionnaires collected during pregnancy and postpartum, and data from the Medical Birth Registry (MBRN) which is a national health registry which holds information on all births in Norway.

*FinnGen*

FinnGen is a national network of Finnish biobanks linked to electronic registry data. It includes data on 500,348 individuals (282,064 females; 12th data release). In this study, we used FinnGen's publicly available genetic association data, since the cohort has both genetic data and pregnancy outcome information available as clinical endpoints (encoded by ICD9-10 codes; see https://www.finngen.fi/en/access_results).

*UK Biobank (UKB)*

The UK Biobank is a cohort study which recruited adults aged 40-69 years registered with the UK National Health Service (NHS) and living within a 25-mile radius of one of the 22 study centres[10]. Participants were invited between 2006 and 2010, with a 5.5% response rate resulting in 500,000 adults being recruited (54.4% female). Here, we used genetic data, self-completed questionnaires, and linked electronic health records including Hospital Episode Statistics (HES) from 1997 for England, 1998 for Wales and 1981 for Scotland; HES also includes maternity-related admissions for England and Wales.

Details on genotyping arrays and imputation for ALSPAC, BiB, MoBa and UKB are provided in the MR-PREG collaboration profile supplementary material[5].

#### Cohort acknowledgments

*Avon Longitudinal Study of Parents and Children (ALSPAC)*
We are extremely grateful to all the families who took part in this study, the midwives for their help in recruiting them, and the whole ALSPAC team, which includes interviewers, computer and laboratory technicians, clerical workers, research scientists, volunteers, managers, receptionists and nurses.

*Born in Bradford (BiB)*
BiB is only possible because of the enthusiasm and commitment of the Children and Parents in BiB. We are grateful to all the participants, teachers, school staff, health professionals and researchers who have made BiB happen.

*Norwegian Mother, Father and Child Cohort Study (MoBa)*
This research has been conducted using MoBa data using application number 2552. MoBa is supported by the Norwegian Ministry of Health and Care services and the Ministry of Education and Research. We are grateful to all the participating families in Norway who take part in this on-going cohort study. We thank the Norwegian Institute of Public Health (NIPH) for generating high-quality genomic data. This research is part of the HARVEST collaboration, supported by the Research Council of Norway (#229624). We also thank the NORMENT Centre for providing genotype data, funded by the Research Council of Norway (#223273), South East Norway Health Authority and KG Jebsen Stiftelsen. We further thank the Center for Diabetes Research, the University of Bergen for providing genotype data and performing QC and imputation of the data funded by the ERC AdG project SELECTionPREDISPOSED, Stiftelsen Kristian Gerhard Jebsen, Trond Mohn Foundation, the Research Council of Norway, the Novo Nordisk Foundation, the University of Bergen, and the Western Norway Health Authorities (Helse Vest).

*FinnGen*
The authors thank the FinnGen investigators for sharing their summary-level data.

*UK Biobank (UKB)*
We would like to thank all the participants of UK Biobank for their vital contribution to the resource. This research has been conducted using the UK Biobank Resource under Application Number 23938.

#### Cohort funding

*Avon Longitudinal Study of Parents and Children (ALSPAC)*
The UK Medical Research Council and Wellcome (Grant ref: 217065/Z/19/Z) and the University of Bristol provide core support for ALSPAC.A comprehensive list of grants funding is available on the ALSPAC website (http://www.bristol.ac.uk/alspac/external/documents/grant-acknowledgements.pdf). ALSPAC GWAS data was generated by Sample Logistics and Genotyping Facilities at Wellcome Sanger Institute and LabCorp (Laboratory Corporation of America) using support from 23andMe. This research was funded in part by the Wellcome Trust (Grant ref: 228276/Z/23/Z)]. For the purpose of Open Access, the author has applied a CC BY public copyright licence to any Author Accepted Manuscript version arising from this submission.

*Born in Bradford (BiB)*
BiB is supported by a Wellcome Longitudinal Population Study Grant (223601/Z/21/Z); a joint grant from the UK Medical Research Council (MRC) and UK Economic and Social Science Research Council (ESRC) (MR/N024391/1); the British Heart Foundation (CS/16/4/32482); a Wellcome Infrastructure Grant (WT101597MA); the National Institute for Health Research under its Applied Research Collaboration for Yorkshire and Humber (NIHR200166). The National Institute for Health Research Clinical Research Network provided research delivery support for this study. The views expressed in this publication are those of the authors and not necessarily those of the National Institute for Health Research or the Department of Health and Social Care.

*Norwegian Mother, Father and Child Cohort Study (MoBa)*
MoBa funding is under **Acknowledgements** as requested by MoBa publication guidelines.

*UK Biobank (UKB)*
UK Biobank is funded primarily by the Wellcome Trust and the Medical Research Council (MRC). It is also funded by the Department of Health, British Heart Foundation, Cancer Research UK, Diabetes UK, National Institute for Health Research (NIHR), Scottish Government, Northwest Regional Development Agency, and Welsh Assembly Government.

#### Cohort ethical approval

*Avon Longitudinal Study of Parents and Children (ALSPAC)*
Ethical approval for the study was obtained from the ALSPAC Ethics and Law Committee and the Local Research Ethics Committees. Consent for biological samples has been collected in accordance with the Human Tissue Act (2004).

Ethical approval was obtained from the ALSPAC Ethics and Law Committee and the Local Research Ethics Committees. Consent for biological samples has been collected in accordance with the Human Tissue Act (2004). Informed consent for the use of data collected via questionnaires and clinics was obtained from participants following the recommendations of the ALSPAC Ethics and Law Committee at the time (details and reference numbers of all ethics approvals can be found at http://www.bristol.ac.uk/medialibrary/sites/alspac/documents/governance/Research%20Ethics%20Committee%20approval%20references.pdf).

*Born in Bradford (BiB)*
Ethical approval for the study was granted by the Bradford National Health Service Research Ethics Committee (ref 06/Q1202/48), and all participants gave written informed consent. The ALL IN sub-study had ethical approval from the London School of Hygiene & Tropical Medicine ethics committee (ref: 5320) and the Bradford Research Ethics committee (ref: 08/H1302/21). Parents (usually the mother) gave informed, written consent to take part in the study.

*Norwegian Mother, Father and Child Cohort Study (MoBa)*
The current study is based on version 12 of the quality-assured data files released for research in 2019. The establishment of MoBa and initial data collection was based on a license from the Norwegian Data Protection Agency and approval from The Regional Committees for Medical and Health Research Ethics. The MoBa cohort is currently regulated by the Norwegian Health Registry Act. The current study was approved by The Regional Committees for Medical and Health Research Ethics of South/East Norway (ref 2018/1256).

*UK Biobank (UKB)*
The UK Biobank has approval from the North West Multi-centre Research Ethics Committee (MREC) as a Research Tissue Bank (RTB) approval. This RTB approval was granted initially in 2011 (11/NW/0382) and it is renewed on a 5-yearly cycle, with the latest one successfully renewed in 2021 (21/NW/0157).

#### References

1. Warrington NM, Beaumont RN, Horikoshi M, Day FR, Helgeland Ø, Laurin C, et al. Maternal and fetal genetic effects on birth weight and their relevance to cardio-metabolic risk factors. Nat Genet. 2019 May;51(5):804–14.

2. Warrington NM, Hwang LD, Nivard MG, Evans DM. Estimating direct and indirect genetic effects on offspring phenotypes using genome-wide summary results data. Nat Commun. 2021 Sept 14;12(1):5420.

3. Wu Y, Zhong X, Lin Y, Zhao Z, Chen J, Zheng B, et al. Estimating genetic nurture with summary statistics of multigenerational genome-wide association studies. Proceedings of the National Academy of Sciences. 2021 June 22;118(25):e2023184118.

4. Beaumont RN, Flatley C, Vaudel M, Wu X, Chen J, Moen GH, et al. Genome-wide association study of placental weight in 179,025 children and parents reveals distinct and shared genetic influences between placental and fetal growth [Internet]. Genetic and Genomic Medicine; 2022 Nov [cited 2023 Dec 15]. Available from: http://medrxiv.org/lookup/doi/10.1101/2022.11.25.22282723

5. McBride N, Clayton GL, Soares ALG, Yang Q, Bond TA, Taylor A, et al. Cohort Profile: The Mendelian Randomization in Pregnancy (MR-PREG) collaboration - Improving evidence for prevention and treatment of adverse pregnancy and perinatal outcomes [Internet]. medRxiv; 2025 [cited 2025 Mar 24]. p. 2025.03.22.25324447. Available from: https://www.medrxiv.org/content/10.1101/2025.03.22.25324447v1

6. Bulik-Sullivan B, Finucane HK, Anttila V, Gusev A, Day FR, Loh PR, et al. An atlas of genetic correlations across human diseases and traits. Nat Genet. 2015 Nov;47(11):1236–41.

7. Boyd A, Golding J, Macleod J, Lawlor DA, Fraser A, Henderson J, et al. Cohort Profile: the 'children of the 90s'--the index offspring of the Avon Longitudinal Study of Parents and Children. Int J Epidemiol. 2013 Feb;42(1):111–27.

8. Fraser A, Macdonald-Wallis C, Tilling K, Boyd A, Golding J, Davey Smith G, et al. Cohort Profile: The Avon Longitudinal Study of Parents and Children: ALSPAC mothers cohort. International Journal of Epidemiology. 2013 Feb 1;42(1):97–110.

9. Wright J, Small N, Raynor P, Tuffnell D, Bhopal R, Cameron N, et al. Cohort Profile: The Born in Bradford multi-ethnic family cohort study. International Journal of Epidemiology. 2013 Aug 1;42(4):978–91.

10. Sudlow C, Gallacher J, Allen N, Beral V, Burton P, Danesh J, et al. UK Biobank: An Open Access Resource for Identifying the Causes of a Wide Range of Complex Diseases of Middle and Old Age. PLOS Medicine. 2015 Mar 31;12(3):e1001779.
